# Burden of Disease from Contaminated Drinking Water in Countries with High Access to Safely Managed Water: A Systematic Review

**DOI:** 10.1101/2022.03.03.22271862

**Authors:** Debbie Lee, Jacqueline MacDonald Gibson, Joe Brown, Jemaneh Habtewold, Heather M. Murphy

## Abstract

The vast majority of residents of high-income countries (≥90%) reportedly have high access to safely managed drinking water. Owing perhaps to the widely held perception of near universal access to high-quality water services in these countries, the burden of waterborne disease in these contexts is understudied. This systematic review aimed to: identify population-scale estimates of waterborne disease in countries with high access to safely managed drinking water, compare methods to quantify disease burden, and identify gaps in available burden estimates. We conducted a systematic review of population-scale disease burden estimates attributed to drinking water in countries where ≥90% of the population has access to safely managed drinking water per official United Nations monitoring. We identified 24 studies reporting estimates for disease burden attributable to microbial contaminants. Across these studies, the population-weighted average burden of gastrointestinal illness risks attributed to drinking water was ∼3,529 annual cases per 100,000 people. Beyond exposure to infectious agents, we identified 10 studies reporting disease burden—predominantly, cancer risks—associated with chemical contaminants. Across these studies, the pooled population-weighted average of excess cancer cases attributable to drinking water was 1.8 annual cancer cases per 100,000 people. These estimates exceed WHO-recommended normative targets for disease burden attributable to drinking water and highlight that there remains important preventable disease burden in these contexts. However, the available literature was scant and limited in geographic scope, disease outcomes, range of microbial and chemical contaminants, and inclusion of subpopulations (rural, low-income communities; Indigenous or Aboriginal peoples; and populations marginalized due to discrimination by race, ethnicity, or socioeconomic status) that could most benefit from water infrastructure investments. Studies quantifying drinking water-associated disease burden in countries with reportedly high access to safe drinking water, with a focus on specific subpopulations and promoting environmental justice, are needed.

## 1. Introduction

Since 1990, the WHO/UNICEF Joint Monitoring Programme for Water Supply, Sanitation and Hygiene has reported on access to safe drinking water worldwide. WHO/UNICEF defines drinking water as safely managed if it comes from an improved source that is accessible on premises, available when needed, and free of fecal and priority chemical contamination. WHO/UNICEF data indicate that in countries classified by the World Bank as low- or middle-income (per-capita incomes <$12,696), major gaps in safe water access remain, with 49.7% of the population unserved by piped water as of 2017 (Deshpande et al., 2020). These gaps manifest in high disease burdens. Recent estimates attribute unsafe drinking water as the cause of 36% of the 1.4 million annual diarrheal deaths worldwide in low- and middle-income countries (Prüss-Ustün et al., 2019).

In contrast to the situation in low- and middle-income countries, WHO/UNICEF data suggest that in high-income countries, most of the population has safe water, with an average of <3% lacking access as of 2017 (Table S1). However, the WHO/UNICEF’s national-scale data and data collection methods can mask local-scale inequities in high-income countries, such as in rural, low-income, and minority communities (Anderson, 2008; Balazs and Ray, 2014). They can also lead to a misconception that water problems of high-income countries have been solved, resulting in underinvestment in maintaining and extending water infrastructure. For example, a recent commentary on infrastructure investment needs in the United States (US) noted “Americans expect the water flowing from their kitchen faucets to be clean and safe,” yet “water infrastructure is particularly vulnerable to neglect” because it “is buried underground or removed from public view, and is thus easily ignored” (Morris, 2017). Given the focus of the global water, sanitation, and hygiene (WASH) sector on low- and middle-income countries, the present work was undertaken to better understand the burden of waterborne disease in high-income countries and to inform a more global understanding of WASH.

Improved understanding of the burden of waterborne disease in countries with high access to safely managed drinking water (most high-income and some middle-income countries) can help identify remaining contamination issues and gaps for water infrastructure investments. To support such efforts, we conducted a systematic review to identify studies estimating the disease burden attributed to contaminated drinking water in the 64 countries/territories with ≥90% safe water access per the WHO/UNICEF definition (Table S1, Figure S1). Among these countries/territories, 54 are classified as high-income; 7 are upper middle income (per-capita incomes between $4,096 and $12,695); and 3 are lower middle income (per-capita incomes between $1,046 and $4,095). The review was intended to:

1. identify available estimates of waterborne disease burden in countries with reportedly high access to safely managed drinking water;
2. evaluate and compare methods to quantify disease burden; and
3. identify gaps in available disease burden estimates.

## 2. Methods

### 2.1 Review Method

We followed the Cochrane Handbook for Systematic Reviews (Higgins et al., 2019) and the Preferred Reporting Items for Systematic reviews and Meta-Analyses (PRISMA) when conducting our review (Page et al., 2021). Table S2 documents study adherence to PRISMA guidelines for reporting.

### 2.2 Search Strategy

We searched PubMed, Web of Science, and SCOPUS for relevant articles published before September 10, 2021. We also searched for grey literature on burden estimates by country on Google and Google Scholar. The objective of this systematic review was to identify articles with explicit population-scale estimates of disease burden, so we limited our search terms accordingly. We searched for articles that included “burden of disease” OR “disease burden” OR “gastrointestinal illness” AND the terms “drinking water” OR “tap water” in text. Last, additional articles were identified from consultations with subject-matter experts and from hand searching reference lists of included articles. Text S1 provides additional details on the development of search criteria.

### 2.3 Inclusion Criteria

Articles in English meeting these inclusion criteria were considered:

a. population-scale burden estimate is for a country where ≥90% of the population has access to safely managed water supplies (as defined by the WHO; Table S1, Figure S1) and
b. estimates cases, hospitalizations, or deaths that could be prevented if ingestion of one or more contaminants in drinking water were prevented.

Articles focusing exclusively on exposure to contaminated water via recreation (e.g., swimming in contaminated water), inhalation, or dermal uptake were excluded. At least two independent reviewers screened the articles against these criteria.

### 2.4 Data Extraction & Synthesis

Disease burden estimates were extracted by co-authors (DL; HM; JMG) using a custom spreadsheet to record estimates, study design, population, disease type(s), contaminant type(s) (microbial or chemical), and drinking water source. Given the limited availability of burden estimates and the wide variation in metrics for characterizing the disease burden, a formal meta-analysis was not possible. However, to enable comparison of estimates across studies, the reported disease burden was converted to incidence per 100,000 population. For some studies, conversion factors were needed to translate the metrics used to report the burden estimate to cases of illness or deaths per 100,000 people. For example, some studies estimated the number of hospitalizations for gastrointestinal illness (GI) attributable to contamination of drinking water. To express these results as total GI cases attributable to drinking water contamination, estimates of the proportion of GI cases resulting in hospitalization were obtained using data for the location where the study was carried out. Table S3 provides details. Disease burden estimates also were compared by country or region. Text S2 outlines approaches for generating these geographic comparisons.

## 3. Results

A total of 514 articles were recovered from the databases, and 14 articles were added by subject-matter experts. After removal of duplicates, 528 articles remained for screening (Figure S2). Thirty-three articles met the inclusion criteria. All but one of the articles focused on a single country or union of countries. Of these 32 articles, most (68.8%) burden estimates were from the United States or Canada; 12.5% were from Asia; 9.4% reported on European countries; and 9.4% were from Australasia (Figure S3, Table S4). Two articles provided estimates from a middle-income country (Iran); the rest focused on high-income countries.

### 3.1 Infectious Diseases

Twenty-four articles estimated GI risks (Table 1). Estimates are difficult to compare across studies because some estimated total GI burden, whereas others quantified the burden only for specific pathogens or specific outcomes (e.g., hospitalization). When sufficient information was available, we converted estimates to units of total cases per 100,000 people to enable cross-study comparisons (Table S3).

**Table 1.**
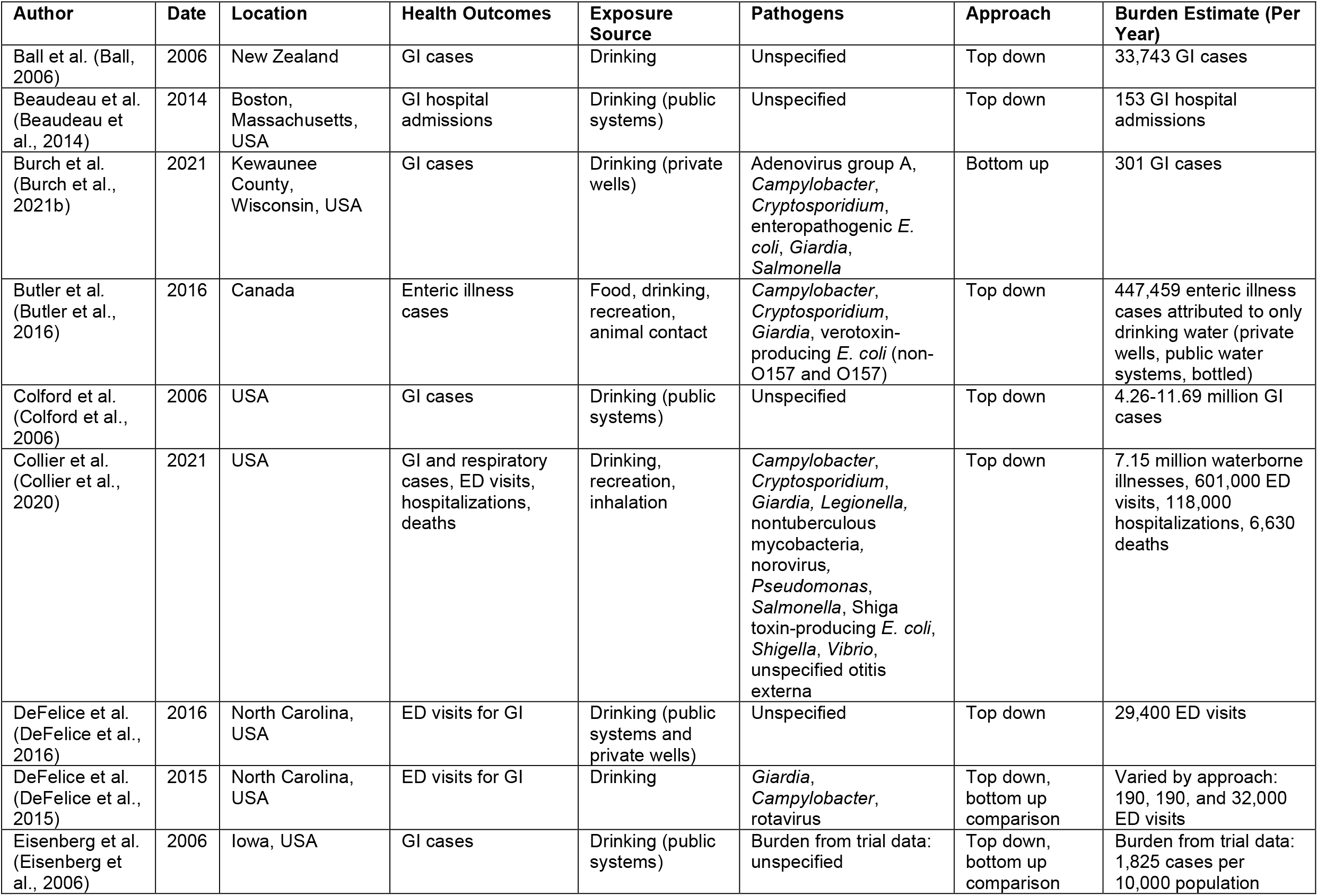
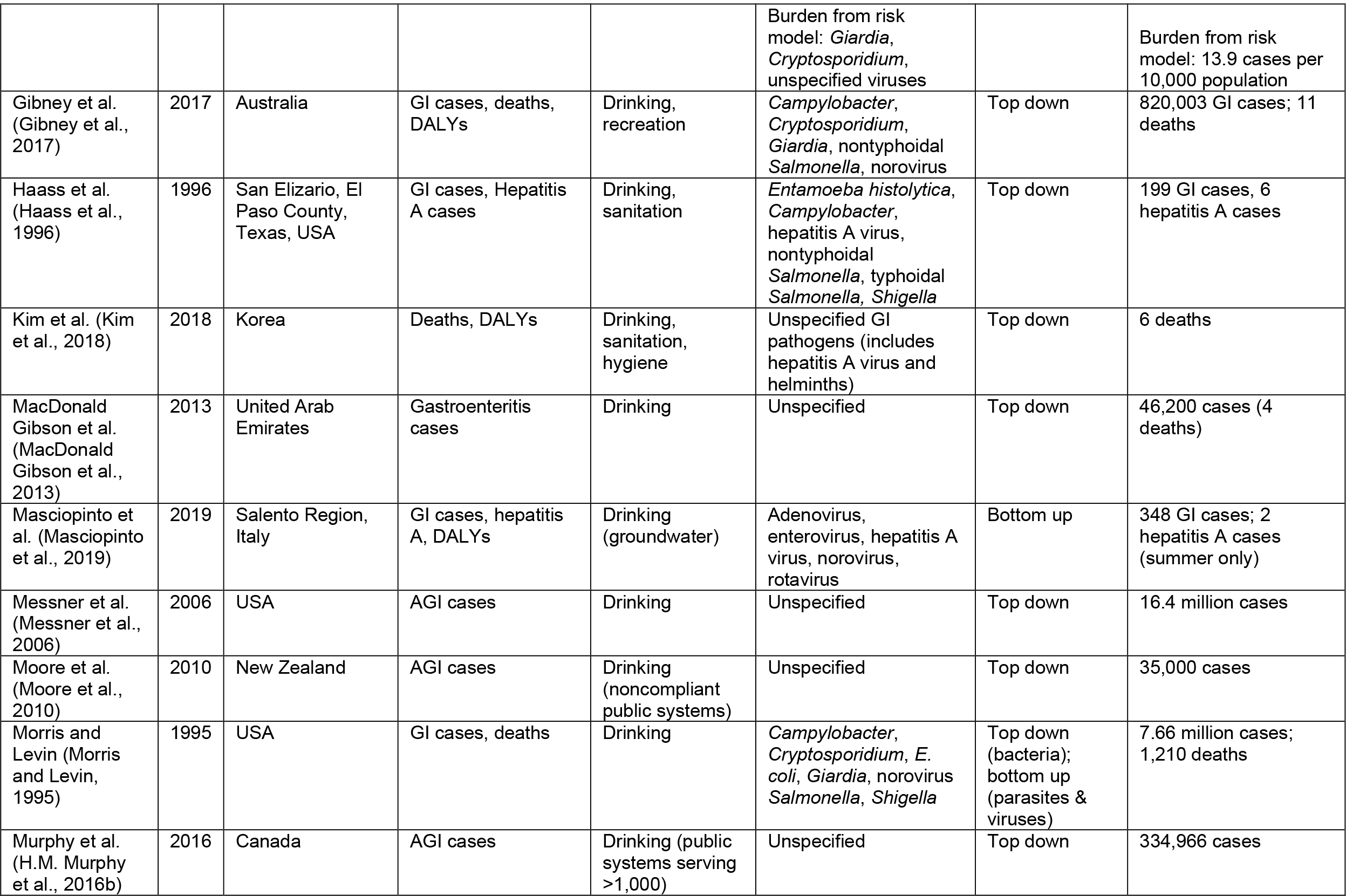
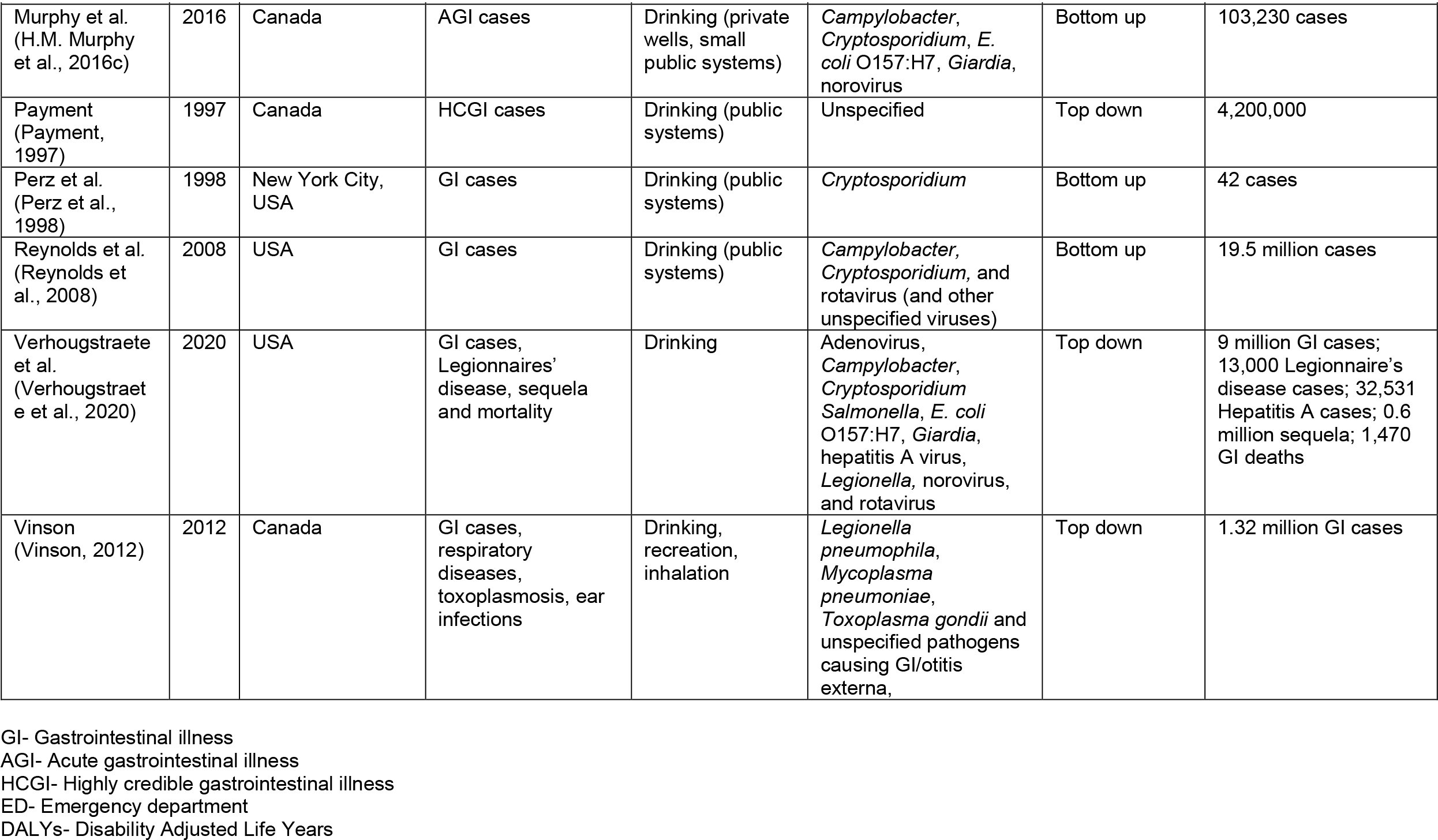
Estimated disease burden from waterborne microbial contaminants in high-income countries

The population-weighted average burden of GI attributable to drinking water was approximately 3,529 cases per 100,000 population (range 40–18,250 per 100,000) across studies that estimated total GI burden or that could be adjusted to estimate this burden. The highest estimate was from a study in Iowa (Eisenberg et al., 2006), using data from a drinking water intervention trial comparing GI cases among participants with and without household water filters (Figure 1). The lowest was from a North Carolina (US) study (DeFelice et al., 2015) that compared three different estimation methods; two resulted in estimates of ∼40 annual cases per 100,000, while the third estimate was ∼6,300 per 100,000 people. Across studies, the population-weighted standard deviation (2,535 per 100,000) was relatively low, just over half the mean.

**Figure 1.**
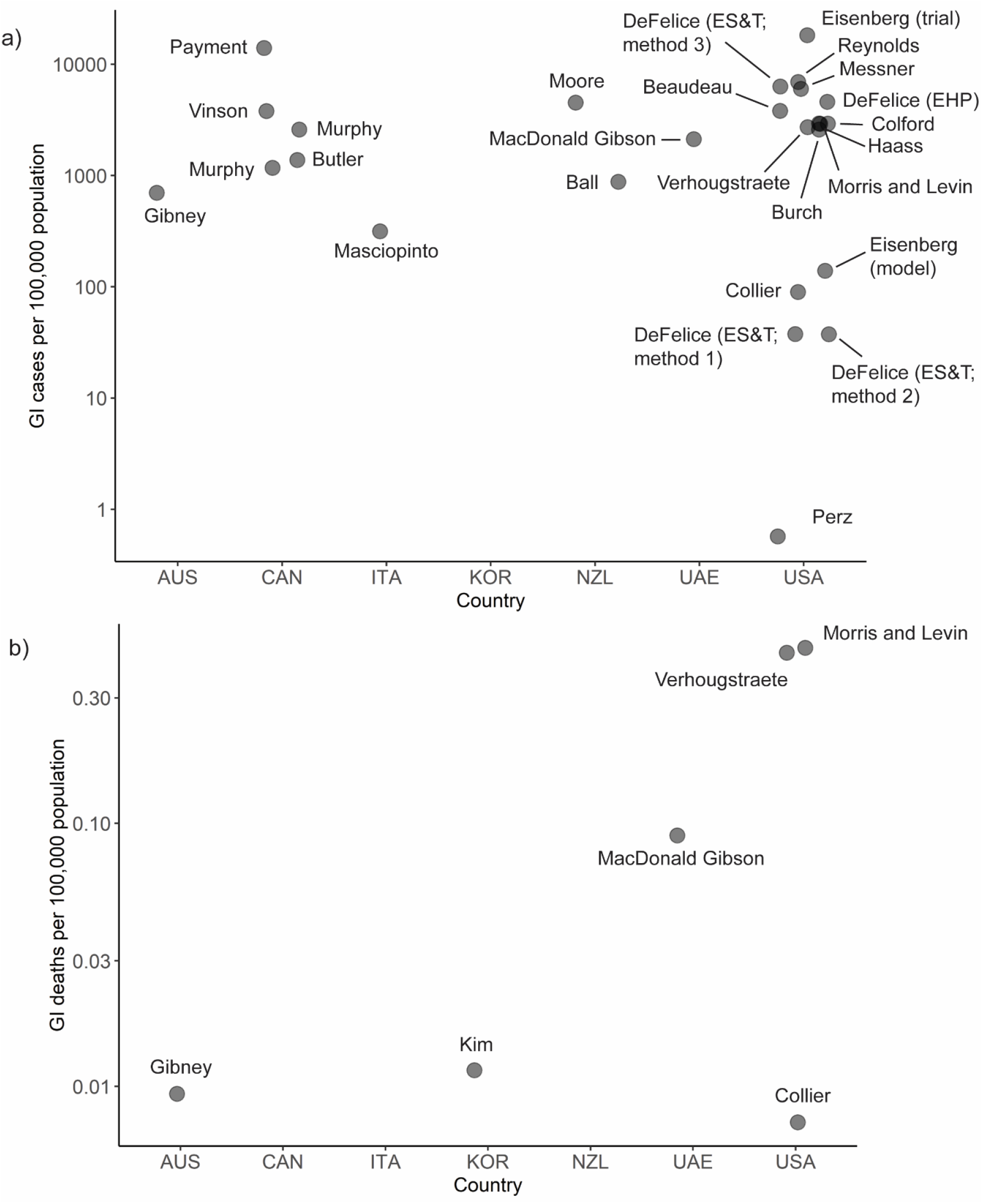
Summary of burden estimates (incidence per 100,000 population) for a) cases of GI illness attributed to microbiological contamination of drinking water and b) deaths from illness attributed to microbiological contamination of drinking water. <u>Countries</u>: AUS=Australia, CAN=Canada, ITA=Italy, KOR=Republic of Korea, NZL=New Zealand, UAE=United Arab Emirates, USA=United States of America

Of the 24 studies, five directly estimated the number of deaths attributable to microbial contaminants. These estimates were very low, averaging 1 per 100,000 people annually (range = 7.3 × 10^−3^ to 2.1 per 100,000).

### 3.2 Non-infectious Diseases

Ten articles estimated the disease burden from chemical contaminants in drinking water (Table 2).

**Table 2.** Estimated disease burden from waterborne chemical contaminants in high-income countries

| Author | Date | Location | Health Outcomes | Exposure Source | Chemical Contaminant | Approach | Burden Estimate (Per Year) |
| --- | --- | --- | --- | --- | --- | --- | --- |
| Abtahi et al. (Abtahi et al., 2019) | 2019 | Iran | Dental fluorosis | Drinking | Fluoride | Bottom up | 60 cases/100,000 people (95% CI 48-69) |
| de Hollander et al. (de Hollander et al., 1999) | 1999 | The Netherlands | Loss of 1-3 IQ points | Drinking | Lead | Bottom up | 1,764 cases |
| DeFelice et al. (DeFelice et al., 2017) | 2017 | North Carolina, USA | Cancer | Drinking (public systems) | 20 carcinogens | Top down | 53.9 (95% CI 27.8-79.4) deaths statewide; individual risk $7.2 \times 10^{-6}$ |
| Dobaradaran et al. (Dobaradaran et al., 2020) | 2020 | Bushehr Province, Iran | Cancer | Drinking, dermal, inhalation | Disinfection byproducts | Bottom up | 1 (95% CI 0.32-2.8) death in Bushehr Province; 95.0 DALYs (94.7–95.2). |
| Evlampidou et al. (Evlampidou et al., 2020) | 2020 | 28 European Union countries | Cancer | Drinking | Disinfection byproducts | Top down | 6,561 (95% CI 3,389, 9,537) cases across the EU |
| Fewtrell et al. (Fewtrell et al., 2006) | 2006 | USA, United Kingdom, New Zealand | Dental and skeletal fluorosis | Drinking | Fluoride | Bottom up | Zero |
| Greco et al. (Greco et al., 2019) | 2019 | USA | Ischemic heart disease; cancer | Drinking (private wells) | Arsenic | Bottom up | 500 ischemic heart disease deaths; 496 fatal cancer cases; 578 nonfatal cancer cases |
| MacDonald Gibson et al. (MacDonald Gibson et al., 2013) | 2013 | United Arab Emirates | Bladder and colorectal cancers | Drinking | Trihalomethanes (THMs) | Top down | 154 healthcare facility visits (and 3 deaths) for bladder cancers and 328 visits (and 9 deaths) for colorectal cancers |
| Mathewson et al. (Mathewson et al., 2020) | 2020 | Wisconsin, USA | Cancer; neural tube defects; low birth weight; preterm birth | Drinking | Nitrate | Top down | 95 (range 46-149 very low birth weight babies; 51 (range 26-79) very preterm births; 1-2 neural tube defect births; 169 (range 111-297) cancer cases |
| Temkin et al. (Temkin et al., 2019) | 2019 | USA | Cancer; neural tube defects; low birth weight; preterm birth | Drinking | Nitrate | Top down | 2,939 very low birth weight cases; 1,725 very preterm births; 41 neural tube defect births; 6,537 (range 2,300-12,594) cancer cases |

Cancers were the most common health outcome considered, included in 7 of the 10 studies (Figure 2). Estimated cancer risks varied depending on the cancer type and contaminants considered. Across all studies with cancer risk estimates, the population-weighted average disease burden was 1.8 excess cases per 100,000 people annually (range = 0.09–2.91).

**Figure 2.**
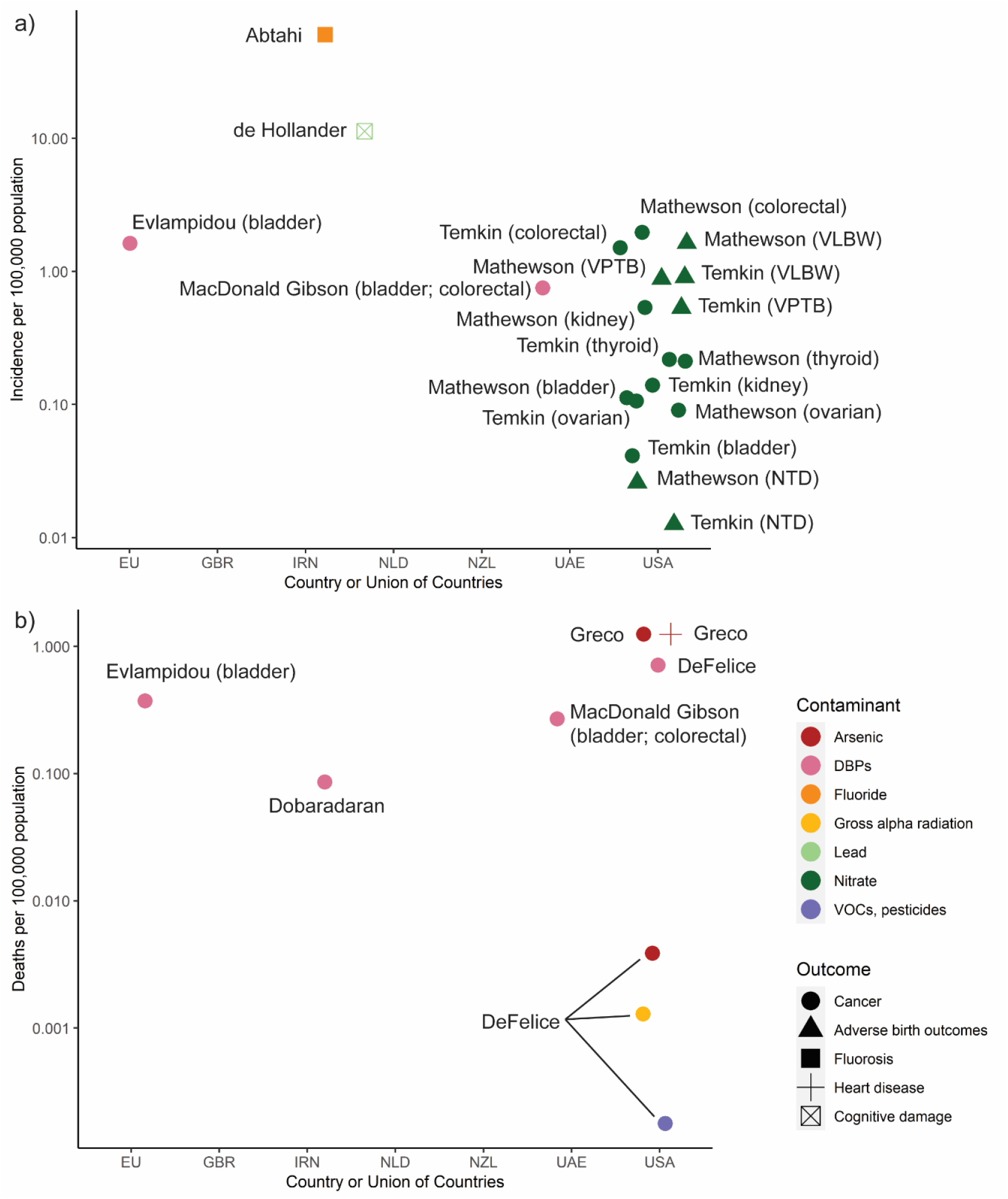
Summary of burden estimates (incidence per 100,000 population) for a) cases of illness attributed to chemical contamination of drinking water, and b) deaths from illness attributed to chemical contamination of drinking water. Note: Estimates of zero cases (per 100,000 population) of fluorosis in USA, GBR (UK), NZL by Fewtrell et al. were excluded <u>Countries</u>: EU=European Union, GBR=United Kingdom, IRN=Iran, NLD=The Netherlands, UAE=United Arab Emirates, USA=United States of America <u>Contaminants</u>: DBPs=Disinfection by-products, VOCs=Volatile Organic Compounds <u>Outcomes</u>: VLBW=Very low birth weight, VPTB=Very preterm birth, NTD=Neural tube defects

Two studies, both in the United States, estimated adverse reproductive outcomes from nitrate in drinking water (Mathewson et al., 2020; Temkin et al., 2019). In both studies, the disease burden was low on a per-capita basis. The highest burden was 1.6 low-birthweight babies per 100,000 people annually.

Surprisingly, despite the recent attention to lead in drinking water resulting from the Flint water crisis (US) and similar events elsewhere, only one study (from the Netherlands) quantified the health burden from lead (de Hollander et al., 1999). That study, published in 1999, estimated that 11 in 100,000 children were losing 1-3 IQ points as a result of neurocognitive damage from lead in drinking water.

The largest reported fatality risks from chemical contaminants were attributed to arsenic and disinfection byproducts (Figure 2b; Figure 3b), though these risks were very low. Across studies, about 0.1–1.3 premature annual deaths per 100,000 people were attributed to these contaminants. A 2017 study from North Carolina (US) attributed fewer than 0.0015 excess annual deaths per 100,000 people to gross alpha radiation, volatile organic compounds, and pesticides in drinking water (DeFelice et al., 2017).

**Figure 3.**
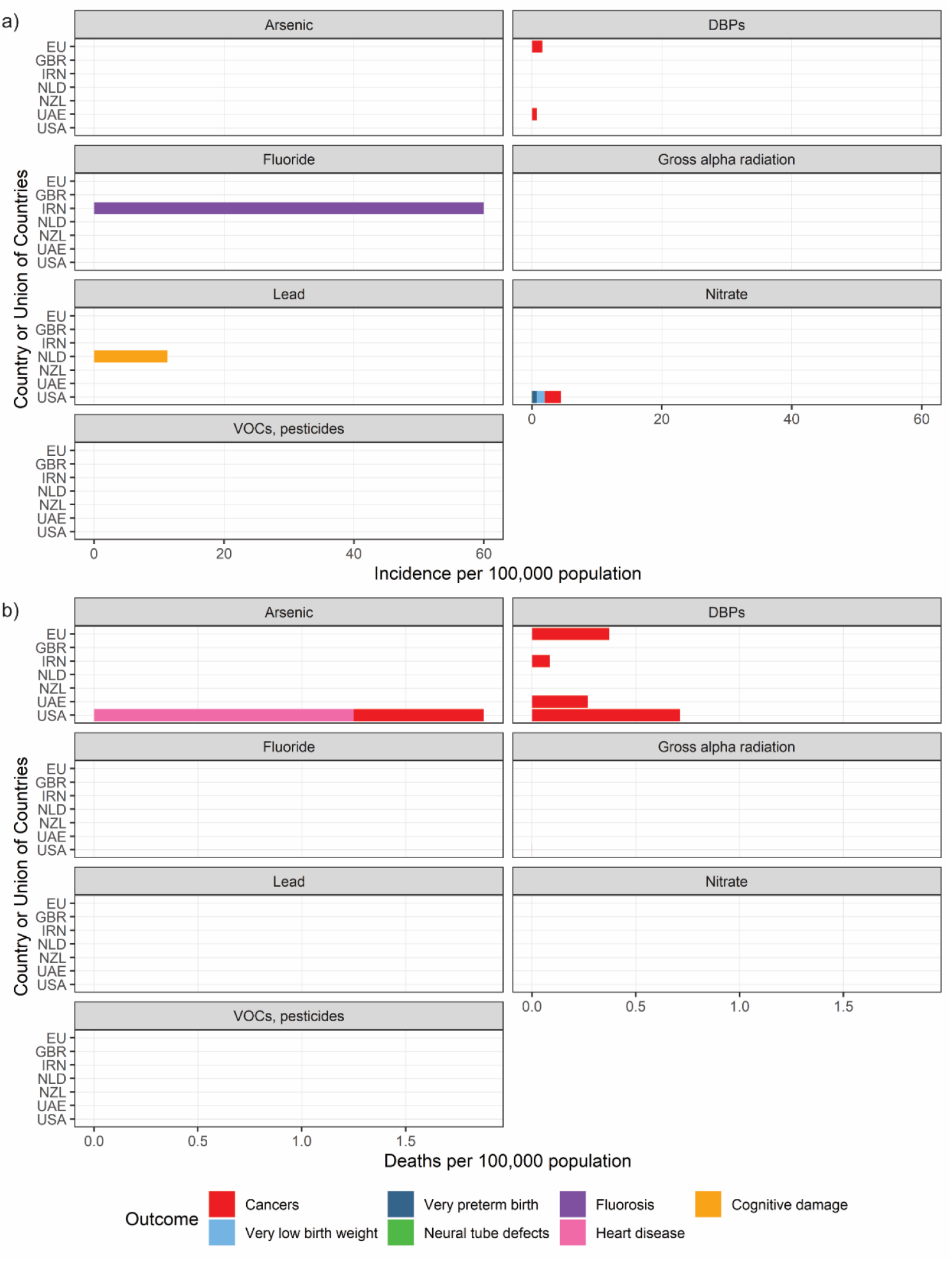
Summary of gaps in the literature on a) cases and b) deaths by chemical contaminant and country. For countries where disease burden estimates were available through multiple studies, disease outcome estimates from different studies were averaged to produce country-specific estimates. For cancers, specific cancer estimates (e.g., bladder cancer, liver cancer) were summed to produce an overall cancer estimate. <u>Note</u>: The burden of neural tube defects in the United States is not visible due to its relatively low level (0.02 cases per 100,000) <u>Countries</u>: EU=European Union, GBR=United Kingdom, IRN=Iran, NLD=The Netherlands, UAE=United Arab Emirates, USA=United States of America <u>Contaminant</u>: DBPs=Disinfection by-products, VOCs=Volatile Organic Compounds

## 4. Discussion

We found 23 studies that estimated the burden of infectious diseases and 9 that estimated the chemical disease burden—and one study that estimated both—from drinking water contamination in countries with reportedly high access to safely managed drinking water. Among the infectious disease burden studies, nearly all focused exclusively on GI; the average burden across studies was approximately 3,529 excess GI cases per 100,000 people yearly (Table 1; Figure 1). Of note, the range in burden estimates was large (40-18,250 cases per 100,000 population), which may be attributed to variability in drinking water sources, study populations, and/or estimation methods. Among studies of non-infectious diseases attributable to chemical contamination, the majority focused on carcinogens. The excess cancer risk across studies averaged 1.8 cases per 100,000 people yearly (ranging from less than one in one billion to two cases per 100,000 people yearly) (Table 2; Figure 2).

These results suggest that risks of GI from drinking water contamination are low in many high-income countries relative to those in developing nations, but that the risks are still higher than recommended targets established by the WHO’s Guidelines for Drinking Water Quality (World Health Organisation, 2017). The Institute of Health Metrics and Evaluation estimates that globally, the burden of disease from unsafe water is 922 annual disability-adjusted life years (DALYs) per 100,000 people, with the highest burden in sub-Saharan Africa, at 2,813 annual DALYs per 100,000 people (“GBD 2019 Resources | Institute for Health Metrics and Evaluation,” n.d.). Using a disability weight of 0.074 for mild diarrhoea as in recent global studies, (Salomon et al., 2015) the population-weighted average 3,529 annual GI cases per 100,000 people equates to approximately 260 annual DALYs per 100,000 people, or less than one-third the global average and about 90% less than in the most affected regions. However, the WHO defines “the tolerable burden of disease … as an upper limit of 10^−6^ DALY per person per year,” equivalent to 0.1 per 100,000 people. By this measure, the burden of infectious diseases associated with drinking water contamination is still much higher than desired, even in high-income nations and even for the lowest risk estimates identified in this review (40 annual GI cases per 100,000 people, equivalent to 3 DALYs per 100,000).

Cancer risk estimates also suggest the magnitude of the disease burden associated with drinking water contamination is relatively low but still exceeds the WHO’s recommendation of one excess annual cancer case per 100,000 people attributed to drinking water. Across studies, the average cancer risk was about twice this amount.

### 3.3 Comparison of Approaches Across Studies

Broadly, study methods can be classified as “top-down” or “bottom up” (Tables 1-2).

#### 3.3.1. Top-Down Approach

Unlike foodborne illness surveillance, a robust system of waterborne illness surveillance does not exist. In the absence of such a system, some studies use overall disease surveillance data. They estimate the fraction of diseases that may be attributable to water and multiply this fraction by the total number of relevant diseases in the population. The WHO uses this method for its periodic global assessments of the contribution of various risk factors to population health (Steenland and Armstrong, 2006).

Top-down studies have used a range of methods to estimate the attributable fraction. Some use previous epidemiologic studies, combined with estimates of the population fraction exposed to various contamination levels. For example, Colford et al. (Colford et al., 2006) and Murphy et al. (H.M. Murphy et al., 2016b) used data from epidemiologic studies in which participants were randomized to receive either an active or sham water treatment system. DeFelice et al. (DeFelice et al., 2017) obtained information on the distribution of occurrence and concentration of all regulated chemical contaminants in public water systems from state and federal water quality surveillance data. Fractions of illnesses attributable to these exposures were estimated by matching exposure concentrations to relative risks of illness from previous epidemiologic studies.

An alternative top-down method, used by Gibney et al. (Gibney et al., 2017), relies on elicitation of expert judgments. Gibney et al. relied on 13 expert opinions on the fractions of illness from five different waterborne pathogens. There was wide variation in expert views. For example, for noroviruses, experts’ median assessments of the attributable fraction ranged from 2% to 50%.

Another top-down approach is population intervention modeling, as illustrated in two studies by DeFelice et al. (DeFelice et al., 2015, 2016). In this approach, disease surveillance data are combined with water source and water quality monitoring data, and a regression model estimating the disease incidence rate as a function of these variables (along with additional controls) is fitted. The number of cases attributable to water contamination is estimated from the regression model by comparing the baseline (observed) rate to the expected rate if water quality were improved.

#### 3.3.2. Bottom-Up Approach

The bottom-up approach combines data on microbial or chemical contaminant exposure concentrations with dose-response information from epidemiologic or toxicologic studies. The dose-response functions are used to predict the estimated probability of illnesses for specific exposure concentrations. Those probabilities are then multiplied by the size of the exposed population. A drawback of this approach is that the estimated number of attributable cases could exceed the total number of actual cases—a potential error that the top-down approach avoids. On the other hand, in the absence of sufficiently comprehensive surveillance data, this is the only alternative (Murphy et al., 2014).

Among studies of the disease burden associated with microbial contamination of drinking water, the bottom-up approach was illustrated by Murphy et al. (H.M. Murphy et al., 2016c), Burch et al. (Burch et al., 2021a), Reynolds et al. (Reynolds et al., 2008), and DeFelice et al. (DeFelice et al., 2015) For example, Murphy et al. used quantitative microbial risk assessment to estimate the disease burden from five pathogens in private wells and small community systems in Canada. Burch *et al*. used well water monitoring data for eight pathogens, along with published dose-response information, to characterize the burden from these pathogens among residents of Kewaunee County, Wisconsin (US), using private wells. Reynolds *et al*. estimated the disease burden attributable to microbial contaminants in US drinking water by making assumptions about the prevalence and concentrations of selected pathogens in different water source types and then using literature-derived dose-response information to estimate illness probabilities from these assumed exposures. DeFelice et al. estimated the burden of GI from three reference pathogens in North Carolina community water systems using monitoring data on the occurrence of *E. coli* and a literature review of the ratio of concentrations of each reference pathogen to *E. coli*.

The bottom-up approach is also common in studies quantifying the disease burden from chemical contaminants. Examples include studies of the disease burden from fluoride (Abtahi et al., 2019) and disinfection byproducts (Dobaradaran et al., 2020) in drinking water in Iran and characterization of the disease burden from arsenic in drinking water in the US (Greco et al., 2019). These studies begin by estimating the probability distribution of contaminant exposure across the study population, and they use a dose-response function derived from previous animal or human studies to predict the number of adverse health outcomes associated with each exposure dose.

Overall, this review indicates that the lack of robust surveillance systems and epidemiological data linking drinking water to adverse health outcomes, particularly for chemical contaminants. The lack of data hinders estimation of the burden of disease attributable to drinking water contamination. A variety of approaches has been used to develop estimates in the face of limited data, but there is no single, agreed-upon approach. This methodological uncertainty complicates comparisons across geographic regions and study years.

### 4.1. Limitations of the Available Studies

This review identified critical gaps in efforts to characterize the disease burden attributable to drinking water contamination in countries with high access to safely managed drinking water. These gaps include a lack of country representation, insufficient spatial resolution in countries with available studies, a limited scope of contaminants considered, and insufficient information to support decision-making about the best ways to decrease the disease burden. Although we attempted to recover estimates from grey literature, only two from New Zealand were found. There may be other unpublished estimates that this review did not recover.

#### 4.1.1. Limited Geographic Representation

The published disease burden studies covered only 10 of 64 countries/territories reporting ≥90% access to safely managed drinking water. One study covered disease burden in the European Union. More than half of studies are from the United States or Canada. As such, the results of this review may be heavily biased toward these countries.

#### 4.1.2. Lack of estimates focusing on specific sub-populations

Even in countries with available disease burden estimates, the estimates are produced at a large scale (i.e., country-wide) and therefore are insufficient to support decision-making about allocation of resources for infrastructure improvements. Such allocation decisions typically begin at the local level and therefore require local-level estimates highlighting populations most in need of assistance. Most available studies provide estimates for large geographic regions (such as an entire country or a large political subdivision within the country), not for smaller spatial units or special subpopulations. The high risks that some populations may face are subsumed when they are averaged in with the general population receiving high-quality water services. Among studies identified in this review, only Haass et al. (Haass et al., 1996) and Beaudeau et al. (Beaudeau et al., 2014) looked at specific vulnerable populations. Haass et al. examined the burden associated with communities in El Paso, Texas (US), inadequately served by water and sanitation services. Beaudeau et al. estimated burden in elderly populations in Massachusetts (US).

As an example, small water systems experience water quality and service delivery problems more often than larger systems due to the lack of economies of scale (Allaire et al., 2018; Cretikos et al., 2009; McFarlane and Harris, 2018). This is of particular concern when considering that small water systems frequently serve rural, low-income communities. Allaire et al. found that violations of health-based drinking water standards in the US often occur in low-income rural areas (Allaire et al., 2018). A cross-sectional study of water quality in small and medium water systems in rural Alabama found an association between self-reported GI symptoms and the following water system characteristics: respondent-reported water supply interruption, low water pressure, lack of total chlorine in water, and detection of *E. coli* in water (Stauber et al., 2016).

Water systems serving communities of color also are more likely to experience water quality problems, with a consequent disease burden that can be masked in studies lacking sufficient spatial resolution. In Canada, boil-water advisories for systems serving Indigenous communities are common—typically a result of inadequate disinfection, mechanical failure, and insufficient technical expertise of system operators (Lui, 2015; Murphy et al., 2015; H.M. Murphy et al., 2016a). A scoping review of drinking water quality and health outcomes of Canadian Indigenous communities found various reports of adverse health outcomes (mostly GI) associated with drinking water but highlighted the paucity of relevant research (Bradford et al., 2016). The remote, Indigenous communities of Australia may also be at higher risk of infection with waterborne pathogens. The water supply in many of these communities often fails to meet microbiological water quality standards (OAG, 2015) and Ng-Hublin et al. found that the notification rate of cryptosporidiosis in Aboriginal people was 50 times that among non-Aboriginal people in Western Australia (Ng-Hublin et al., 2017). Schaider et al. found a positive association across the United States between the proportion of Hispanic residents and nitrate levels in drinking water (Schaider et al., 2019). Balasz et al. found a similar association in the San Joaquin Valley of California for small community water systems serving a large proportion of Latino residents (Balazs et al., 2011). A study by Nigra and Navas-Acien found that incarcerated people in the southwestern US were disproportionately at risk of exposure to arsenic in drinking water (Nigra and Navas-Acien, 2020). MacDonald Gibson et al. found that in Wake County, North Carolina (US), children in households relying on private wells had higher blood lead levels than those with community water service and that these differences were especially acute in peri-urban minority communities (MacDonald Gibson et al., 2020).

#### 4.1.3. Limited Scope of Disease Types and Contaminants

Nearly all the studies of the waterborne infectious disease burden consider only GI, overlooking other critical diseases (such as respiratory illnesses) that can be transmitted through drinking water. For example, only three studies identified in this review (Collier et al., 2020; Verhougstraete et al., 2020; Vinson, 2012) characterized the respiratory disease burden attributable to *Legionella*, even though it caused all deaths from waterborne disease outbreaks in the United States in 2013–2014 (Benedict et al., 2017).

More information on the comparative disease burden from chemical contaminants is especially important for developed nations since these contaminants tend to be the major focus of drinking water regulations in developed nations (DeFelice et al., 2017; Roberson, 2011). Especially notable is the dearth of studies estimating the disease burden from lead in drinking water, despite recent water crises brought about by lead (for example, in Flint, Michigan, US), along with mounting evidence that exposure to lead in private well water may be much more prevalent than previously recognized and, in some cases, can be a source of health disparities (Hanna-Attisha et al., 2016; MacDonald Gibson et al., 2020). This review located only one study that quantified the burden of disease due to lead in drinking water in a developed nation, and it was completed more than 20 years ago. This finding is surprising, since lead is ubiquitous in plumbing (even in plumbing branded as “lead-free”) (Katner et al., 2016). Updated estimates of lead exposure risks from drinking water and the associated disease burden are needed across developed nations.

Also needed are studies of the disease burden attributable to “emerging” chemical contaminants—that is, those for which knowledge on their potential prevalence in drinking water is relatively new. One example is per- and polyfluoroalkyl substances (PFAS), which have been reported in drinking water sources and systems globally in recent years (Domingo and Nadal, 2019). Another example is toxins formed by aquatic fungi and algae, such as mycotoxins and cyanotoxins (Székács, 2021). The most recently published waterborne disease surveillance data from the US Centers for Disease Control and Prevention indicate that cyanotoxins caused 12% of reported US waterborne disease cases (Benedict et al., 2017).

#### 4.1.4. Insufficient Consideration of Causal Factors

Also lacking is information that would enable estimates of specific water system deficiencies causing preventable illness. Available estimates focus on total disease burden, not on contributing factors. These might include source water contamination, treatment system deficiencies or failures, distribution system breaches, disinfectant loss in the distribution system, water shutoffs and low-pressure events, or release or formation of contaminants as water travels through plumbing within buildings.

#### 4.1.5. Strengths and Limitations

This review is a first attempt to summarize the global evidence for waterborne microbial and chemical disease burdens in high-income countries. Current estimates (Vos et al., 2020) using alternative methods have assumed substantially lower attributable burdens where water infrastructure coverage is high. Our review shows that these estimates are likely to be too conservative. Where burden estimates exist, they are typically much higher, even for the very limited range of contaminants that have been the focus of burden estimates. Our review additionally reveals that there are major gaps in burden estimation for high-income countries, suggesting that it is a lack of data – not lack of burden – that has led to a perception that economically advanced countries no longer face waterborne diseases. This perception may lead to underinvestment in infrastructure.

Despite our review providing a useful summary of evidence, our analysis comes with clear limitations. First, we were limited by our reliance on the WHO/UNICEF definition of safely managed drinking water. These estimates of the proportion of the population with safely managed water may not be accurate, though they are the only global estimates of infrastructure coverage and are widely used to track progress in meeting the Sustainable Development Goals. As one example, according to the WHO/UNICEF, 99% of the US population has access to safely managed drinking water but nearly 13% of the US population relies on federally unregulated private wells for drinking water (Dieter et al., 2018). Households in the US are responsible for monitoring and maintaining their own wells, and thus, the safety of these water supplies remains unknown. One major issue identified by this review was the disparity between the perception of near universal access to safely managed drinking water in high-income countries (as indicated by the WHO/UNICEF definition) and the blind spots in our understanding of water quality for a sizeable proportion of the population within these high-income countries. This disconnect may be perpetuating the information gaps about the burden of disease attributed to drinking water in these contexts.

Second, this review focused on areas where ≥90% of the population had access to safely managed drinking water, which meant that, up to 10% of the population in countries/territories included in the review did not have access to safely managed drinking water. Many of the burden estimates did not differentiate by water source (surface water vs. groundwater) or system (public vs. private) while others did. Comparisons of burden are complicated by the potential variations in water quality within a country depending on drinking water source. This further highlights the need for higher resolution disease burden data for high-income countries.

A third limitation includes the low number of available studies, and their narrow range of contaminants and the limited diversity of their methods, which makes synthesis impossible. We also limited our review to studies in English, which may have eliminated burden estimates from many countries.

Limitations notwithstanding, we conducted a rigorous systematic review of the literature to identify and compare estimates of burden attributed to drinking water contamination in countries perceived to provide near universal access to safely managed drinking water. The major finding and takeaway of this review is precisely the lack of burden estimates available. The dearth of these estimates in countries with robust water infrastructure coupled with the evidence of water quality issues for subpopulations of these countries and the limitations of relying on WHO/UNICEF definitions of safely managed drinking water may suggest that the true burden of disease attributed to drinking water contamination may be greater in these countries than previously recognized.

## 5. Conclusions

This review sheds light on the dearth of research on the disease burden attributable to unsafe drinking water in countries with reportedly high access to safe drinking water. Estimating this disease burden remains difficult given the lack of a comprehensive surveillance system and gaps in drinking water quality data. WHO/UNICEF data suggesting the vast majority of the population has access to safely managed water can be misleading because these data mask variation in the quality of water service delivery that can have important public health consequences. Further research on the drinking water-attributed disease burden in these countries remains vital to identifying the main drivers of waterborne disease, the populations at greatest risk, and strategies for improving health outcomes related to drinking water.

## Data Availability

Extracted data are available on request to the corresponding author.

## Contributors

HMM, JMG, and JB designed the study. DL, JH, JMG, and HMM recovered articles, assessed article eligibility, and extracted and analyzed data from articles. DL, JMG, and HMM wrote the initial drafts of the manuscript. DL, JH, JMG, HMM, and JB contributed to edits of the manuscript.

## Data sharing

Extracted data are available on request to the corresponding author.

## Acknowledgements

We would like to thank Jessica Evans, Ilya Law (IL), Tala Alahdab (TA) for their help screening titles and abstracts for this review.

## Protocol Registration

A protocol for this review was not formally registered.

## References

Abtahi, M., Dobaradaran, S., Jorfi, S., Koolivand, A., Khaloo, S.S., Spitz, J., Saeedi, H., Golchinpour, N., Saeedi, R., 2019. Age-sex specific disability-adjusted life years (DALYs) attributable to elevated levels of fluoride in drinking water: A national and subnational study in Iran, 2017. WATER Res. 157, 94–105. https://doi.org/10.1016/j.watres.2019.03.087

Allaire, M., Wu, H., Lall, U., 2018. National trends in drinking water quality violations. Proc. Natl. Acad. Sci. U. S. A. 115, 2078–2083. https://doi.org/10.1073/pnas.1719805115

Anderson, M.W., 2008. Cities inside out: race, poverty, and exclusion at the urban fringe. UCLA Law Rev. 55, 1095–1160. https://doi.org/10.1111/j.1467-8276.2009.01305.x

Balazs, C., Morello-Frosch, R., Hubbard, A., Ray, I., 2011. Social disparities in nitrate-contaminated drinking water in California’s San Joaquin Valley. Environ. Health Perspect. 119, 1272–1278. https://doi.org/10.1289/ehp.1002878

Balazs, C.L., Ray, I., 2014. The drinking water disparities framework: on the origins and persistence of inequities in exposure. Am. J. Public Health 104, 603–611. https://doi.org/10.2105/AJPH.2013.301664

Ball, A., 2006. Estimation of the burden of water-borne disease in New Zealand: preliminary report. Christchurch Environ. Sci. Res. Ltd.

Beaudeau, P., Schwartz, J., Levin, R., 2014. Drinking water quality. and hospital admissions of elderly people for gastrointestinal illness in Eastern Massachusetts, 1998-2008. WATER Res. 52, 188–198. https://doi.org/10.1016/j.watres.2014.01.005

Benedict, K.M., Reses, H., Vigar, M., Roth, D.M., Roberts, V.A., Mattioli, M., Cooley, L.A., Hilborn, E.D., Wade, T.J., Fullerton, K.E., Yoder, J.S., Hill, V.R., 2017. Surveillance for Waterborne Disease Outbreaks Associated with Drinking Water — United States, 2013–2014. MMWR. Morb. Mortal. Wkly. Rep. 66, 1216–1221. https://doi.org/10.15585/mmwr.mm6644a3

Bradford, L.E.A., Bharadwaj, L.A., Okpalauwaekwe, U., Waldner, C.L., 2016. Drinking water quality in Indigenous communities in Canada and health outcomes: a scoping review. Int. J. Circumpolar Health 75. https://doi.org/10.3402/ijch.v75.32336

Burch, T.R., Stokdyk, J.P., Spencer, S.K., Kieke, B.A., Firnstahl, A.D., Muldoon, M.A., Borchardt, M.A., 2021a. Quantitative Microbial Risk Assessment for Contaminated Private Wells in the Fractured Dolomite Aquifer of Kewaunee County, Wisconsin. Environ. Health Perspect. 129. https://doi.org/10.1289/EHP7815

Burch, T.R., Stokdyk, J.P., Spencer, S.K., Kieke, B.A., Firnstahl, A.D., Muldoon, M.A., Borchardt, M.A., 2021b. Quantitative Microbial Risk Assessment for Contaminated Private Wells in the Fractured Dolomite Aquifer of Kewaunee County, Wisconsin. Environ. Health Perspect. 129. https://doi.org/10.1289/EHP7815

Butler, A.J., Pintar, K.D.M., Thomas, M.K., 2016. Estimating the Relative Role of Various Subcategories of Food, Water, and Animal Contact Transmission of 28 Enteric Diseases in Canada. Foodborne Pathog. Dis. 13, 57–64. https://doi.org/10.1089/fpd.2015.1957

Colford, J.M., Roy, S., Beach, M.J., Hightower, A., Shaw, S.E., Wade, T.J., 2006. A review of household drinking water intervention trials and an approach to the estimation of endemic waterborne gastroenteritis in the United States. J. Water Health 4, 71–88.

Collier, S.A., Deng, L., Adam, E.A., Benedict, K.M., Beshearse, E.M., Blackstock, A.J., Bruce, B.B., Derado, G., Edens, C., Fullerton, K.E., 2020. Estimate of burden and direct healthcare cost of infectious waterborne disease in the United States.

Cretikos, M., Byleveld, P., Durrheim, D.N., Porigneaux, P., Merritt, T., Leask, S., 2009. Supply system factors associated with microbiological drinking water safety in regional New South Wales, Australia, 2001–2007. J. Water Health 8, 257–268. https://doi.org/10.2166/wh.2009.203

de Hollander, A.E.M., Melse, J.M., Lebret, E., Kramers, P.G.N., 1999. An aggregate public health indicator to represent the impact of multiple environmental exposures. Epidemiology 10, 606–617. https://doi.org/10.1097/00001648-199909000-00030

DeFelice, N.B., Johnston, J.E., MacDonald Gibson, J., 2016. Reducing Emergency Department Visits for Acute Gastrointestinal Illnesses in North Carolina (USA) by Extending Community Water Service. Environ. Health Perspect. 124, 1583–1591. https://doi.org/10.1289/EHP160

DeFelice, N.B., Johnston, J.E., MacDonald Gibson, J., 2015. Acute gastrointestinal illness risks in North Carolina community water Systems: a methodological comparison. Environ. Sci. Technol. 49, 10019–10027. https://doi.org/10.1021/acs.est.5b01898

DeFelice, N.B., Leker, H.G., MacDonald Gibson, J., 2017. Annual cancer risks from chemicals in North Carolina community water systems. Hum. Ecol. Risk Assess. 23, 974–991. https://doi.org/10.1080/10807039.2017.1292842

Deshpande, A., Miller-Petrie, M.K., Lindstedt, P.A., Baumann, M.M., Johnson, K.B., Blacker, B.F., Abbastabar, H., Abd-Allah, F., Abdelalim, A., Abdollahpour, I., 2020. Mapping geographical inequalities in access to drinking water and sanitation facilities in low-income and middle-income countries, 2000–17. Lancet Glob. Heal. 8, e1162–e1185.

Dieter, C.A., Maupin, M.A., Caldwell, R.R., Harris, M.A., Ivahnenko, T.I., Lovelace, J.K., Barber, N.L., Linsey, K.S., 2018. Estimated use of water in the United States in 2015, Circular. Reston, VA. https://doi.org/10.3133/cir1441

Dobaradaran, S., Fard, E.S., Tekle-Rottering, A., Keshtkar, M., Karbasdehi, V.N., Abtahi, M., Gholamnia, R., Saeedi, R., 2020. Age-sex specific and cause-specific health risk and burden of disease induced by exposure to trihalomethanes (THMs) and haloacetic acids (HAAs) from drinking water: An assessment in four urban communities of Bushehr Province, Iran, 2017. Environ. Res. 182. https://doi.org/10.1016/j.envres.2019.109062

Domingo, J.L., Nadal, M., 2019. Human exposure to per- and polyfluoroalkyl substances (PFAS) through drinking water: A review of the recent scientific literature. Environ. Res. 177, 108648. https://doi.org/https://doi.org/10.1016/j.envres.2019.108648

Eisenberg, J.N.S., Hubbard, A., Wade, T.J., Sylvester, M.F., LeChevallier, M.W., Levy, D.A., Colford, J.M., 2006. Inferences Drawn from a Risk Assessment Compared Directly with a Randomized Trial of a Home Drinking Water Intervention. Environ. Health Perspect. 114, 1199–1204. https://doi.org/10.1289/ehp.8682

Evlampidou, I., Font-Ribera, L., Rojas-Rueda, D., Gracia-Lavedan, E., Costet, N., Pearce, N., Vineis, P., Jaakkola, J.J.K., Delloye, F., Makris, K.C., Stephanou, E.G., Kargaki, S., Kozisek, F., Sigsgaard, T., Hansen, B., Schullehner, J., Nahkur, R., Galey, C., Zwiener, C., Vargha, M., Righi, E., Aggazzotti, G., Kalnina, G., Grazuleviciene, R., Polanska, K., Gubkova, D., Bitenc, K., Goslan, E.H., Kogevinas, M., Villanueva, C.M., 2020. Trihalomethanes in Drinking Water and Bladder Cancer Burden in the European Union. Environ. Health Perspect. 128. https://doi.org/10.1289/EHP4495

Fewtrell, L., Smith, S., Kay, D., Bartram, J., 2006. An attempt to estimate the global burden of disease due to fluoride in drinking water. J. Water Health 4, 533–542.

GBD 2019 Resources | Institute for Health Metrics and Evaluation [WWW Document], n.d.

Gibney, K.B., O’Toole, J., Sinclair, M., Leder, K., 2017. Burden of Disease Attributed to Waterborne Transmission of Selected Enteric Pathogens, Australia, 2010. Am. J. Trop. Med. Hyg. 96, 1400–1403. https://doi.org/10.4269/ajtmh.16-0907

Greco, S.L., Belova, A., Haskell, J., Backer, L., 2019. Estimated burden of disease from arsenic in drinking water supplied by domestic wells in the United States. J. Water Health 17, 801–812. https://doi.org/10.2166/wh.2019.216

Haass, J.A., Miller, G.L., Haddix, A.C., Nickey, L.N., Sinks, T., 1996. An economic analysis of water and sanitation infrastructure improvements in the Colonias of EI Paso County, Texas. Int. J. Occup. Environ. Health 2, 211–221.

Hanna-Attisha, M., LaChance, J., Sadler, R.C., Schnepp, A.C., 2016. Elevated blood lead levels in children associated with the Flint drinking water crisis: A spatial analysis of risk and public health response. Am. J. Public Health 106, 283–290. https://doi.org/10.2105/AJPH.2015.303003

Higgins, J., Thomas, J., Chandler, J., Cumpston, M., Li, T., Page, M., Welch, V., 2019. Cochrane Handbook for Systematic Reviews of Interventions, 2nd ed. John Wiley & Sons, Chichester, UK.

Katner, A., Pieper, K.J., Lambrinidou, Y., Brown, K., Hu, C.-Y., Mielke, H.W., Edwards, M.A., 2016. Weaknesses in Federal Drinking Water Regulations and Public Health Policies that Impede Lead Poisoning Prevention and Environmental Justice. Environ. Justice 9, 109–117. https://doi.org/10.1089/env.2016.0012

Kim, J.H., Cheong, H.K., Jeon, B.H., 2018. Burden of Disease Attributable to Inadequate Drinking Water, Sanitation, and Hygiene in Korea. J. KOREAN Med. Sci. 33. https://doi.org/10.3346/jkms.2018.33.e288

Lui, E., 2015. On notice for a drinking water crisis in Canada. Council of Canadians Ottawa.

MacDonald Gibson, J., Fisher, M., Clonch, A., MacDonald, J., Cook, P., 2020. Children drinking private well water have higher blood lead than those with city water. Proc. Natl. Acad. Sci. 117, 16898–16907. https://doi.org/10.1073/pnas.2002729117

MacDonald Gibson, J., Thomsen, J., Launay, F., Harder, E., DeFelice, N.B., 2013. Deaths and medical visits attributable to environmental pollution in the United Arab Emirates. PLoS One 8, e57536–e57536. https://doi.org/10.1371/journal.pone.0057536

Masciopinto, C., De Giglio, O., Scrascia, M., Fortunato, F., La Rosa, G., Suffredini, E., Pazzani, C., Prato, R., Montagna, M.T., 2019. Human health risk assessment for the occurrence of enteric viruses in drinking water from wells: Role of flood runoff injections. Sci. Total Environ. 666, 559–571. https://doi.org/10.1016/j.scitotenv.2019.02.107

Mathewson, P.D., Evans, S., Byrnes, T., Joos, A., Naidenko, O. V, 2020. Health and economic impact of nitrate pollution in drinking water: a Wisconsin case study. Environ. Monit. Assess. 192, 1–18.

McFarlane, K., Harris, L.M., 2018. Small systems, big challenges: review of small drinking water system governance. Environ. Rev. 26, 378+.

Messner, M., Shaw, S., Regli, S., Rotert, K., Blank, V., Soller, J., 2006. An approach for developing a national estimate of waterborne disease due to drinking water and a national estimate model application. J. Water Health 4, 201–240.

Moore, D., Black, M., Valji, Y., Tooth, R., 2010. Cost Benefit Analysis of Raising the Quality of New Zealand Networked Drinking Water.

Morris, J.C., 2017. Planning for Water Infrastructure: Challenges and Opportunities. Public Work. Manag. Policy 22, 24–30. https://doi.org/10.1177/1087724X16668182

Morris, R.D., Levin, R., 1995. Estimating the incidence of waterborne infectious disease related to drinking water in the United States. IAHS Publ. Proc. Reports-Intern Assoc Hydrol. Sci. 233, 75–88.

Murphy, H.M., Bhatti, M., Harvey, R., McBean, E.A., 2016a. Using Decision Trees to Predict Drinking Water Advisories in Small Water Systems. J. AWWA 108, E109–E118. https://doi.org/https://doi.org/10.5942/jawwa.2016.108.0008

Murphy, H.M., Corston-Pine, E., Post, Y., McBean, E.A., 2015. Insights and opportunities: challenges of Canadian First Nations drinking water operators. Int. Indig. Policy J. 6.

Murphy, H.M., Pintar, K.D.M., McBean, E.A., Thomas, M.K., 2014. A systematic review of waterborne disease burden methodologies from developed countries. J. Water Health 12, 634–655. https://doi.org/http://dx.doi.org/10.2166/wh.2014.049

Murphy, H.M., Thomas, M.K., Medeiros, D.T., McFadyen, S., Pintar, K.D.M., 2016b. Estimating the number of cases of acute gastrointestinal illness (AGI) associated with Canadian municipal drinking water systems. Epidemiol. Infect. 144, 1371–1385. https://doi.org/10.1017/S0950268815002083

Murphy, H.M., Thomas, M.K., Schmidt, P.J., Medeiros, D.T., McFadyen, S., Pintar, K.D.M., 2016c. Estimating the burden of acute gastrointestinal illness due to Giardia, Cryptosporidium, Campylobacter, E. coli O157 and norovirus associated with private wells and small water systems in Canada. Epidemiol. Infect. 144, 1355–1370. https://doi.org/10.1017/S0950268815002071

Ng-Hublin, J.S.Y., Combs, B., Reid, S., Ryan, U., 2017. Differences in the occurrence and epidemiology of cryptosporidiosis in Aboriginal and non-Aboriginal people in Western Australia (2002-2012). Infect. Genet. Evol. 53, 100–106. https://doi.org/https://doi.org/10.1016/j.meegid.2017.05.018

Nigra, A.E., Navas-Acien, A., 2020. Arsenic in US correctional facility drinking water, 2006–2011. Environ. Res. 188, 109768. https://doi.org/https://doi.org/10.1016/j.envres.2020.109768

OAG, 2015. Delivering Essential Services to Remote Aboriginal Communities.

Page, M.J., McKenzie, J.E., Bossuyt, P.M., Boutron, I., Hoffmann, T.C., Mulrow, C.D., Shamseer, L., Tetzlaff, J.M., Akl, E.A., Brennan, S.E., Chou, R., Glanville, J., Grimshaw, J.M., Hróbjartsson, A., Lalu, M.M., Li, T., Loder, E.W., Mayo-Wilson, E., McDonald, S., McGuinness, L.A., Stewart, L.A., Thomas, J., Tricco, A.C., Welch, V.A., Whiting, P., Moher, D., 2021. The PRISMA 2020 statement: An updated guideline for reporting systematic reviews. BMJ 372. https://doi.org/10.1136/BMJ.N71

Payment, P., 1997. Epidemiology of endemic gastrointestinal and respiratory diseases: Incidence, fraction attributable to tap water and costs to society. WATER Sci. Technol. 35, 7–10. https://doi.org/10.1016/S0273-1223(97)00226-6

Perz, J.F., Ennever, F.K., Le Blancq, S.M., 1998. Cryptosporidium in Tap Water: Comparison of Predicted Risks with Observed Levels of Disease. Am. J. Epidemiol. 147, 289–301. https://doi.org/10.1093/oxfordjournals.aje.a009449

Prüss-Ustün, A., Wolf, J., Bartram, J., Clasen, T., Cumming, O., Freeman, M.C., Gordon, B., Hunter, P.R., Medlicott, K., Johnston, R., 2019. Burden of disease from inadequate water, sanitation and hygiene for selected adverse health outcomes: An updated analysis with a focus on low- and middle-income countries. Int. J. Hyg. Environ. Health 222, 765–777. https://doi.org/10.1016/j.ijheh.2019.05.004

Reynolds, K., D Mena K., Gerba, C., 2008. Risk of Waterborne Illness Via Drinking Water in the United States, Reviews of environmental contamination and toxicology. https://doi.org/10.1007/978-0-387-71724-1_4

Roberson, J.A., 2011. What’s next after 40 years of drinking water regulations? Environ. Sci. Technol. 45, 154–60. https://doi.org/10.1021/es101410v

Salomon, J.A., Haagsma, J.A., Davis, A., de Noordhout, C.M., Polinder, S., Havelaar, A.H., Cassini, A., Devleesschauwer, B., Kretzschmar, M., Speybroeck, N., Murray, C.J.L., Vos, T., 2015. Disability weights for the Global Burden of Disease 2013 study. Lancet Glob. Heal. 3, e712–e723. https://doi.org/10.1016/S2214-109X(15)00069-8

Schaider, L.A., Swetschinski, L., Campbell, C., Rudel, R.A., 2019. Environmental justice and drinking water quality: are there socioeconomic disparities in nitrate levels in U.S. drinking water? Environ. Heal. 18, 3. https://doi.org/10.1186/s12940-018-0442-6

Stauber, C.E., Wedgworth, J.C., Johnson, P., Olson, J.B., Ayers, T., Elliott, M., Brown, J., 2016. Associations between Self-Reported Gastrointestinal Illness and Water System Characteristics in Community Water Supplies in Rural Alabama: A Cross-Sectional Study. PLoS One 11. https://doi.org/10.1371/journal.pone.0148102

Steenland, K., Armstrong, B., 2006. An overview of methods for calculating the burden of disease due to specific risk factors. Epidemiology 17, 512–9. https://doi.org/10.1097/01.ede.0000229155.05644.43

Székács, A., 2021. Mycotoxins as emerging contaminants. Introduction to the special issue “rapid detection of mycotoxin contamination.” Toxins (Basel). https://doi.org/10.3390/toxins13070475

Temkin, A., Evans, S., Manidis, T., Campbell, C., Naidenko, O. V, 2019. Exposure-based assessment and economic valuation of adverse birth outcomes and cancer risk due to nitrate in United States drinking water. Environ. Res. 176, 108442. https://doi.org/https://doi.org/10.1016/j.envres.2019.04.009

Verhougstraete, M., Reynolds, K.A., Pearce-Walker, J., Gerba, C., 2020. Cost-benefit analysis of point-of-use devices for health risks reduction from pathogens in drinking water. J. Water Health 18, 968–982. https://doi.org/10.2166/wh.2020.111

Vinson, N.G., 2012. Towards estimating the economic burden of waterborne illness in Canada: what do we know, where do we go, in: Poster Presented at The Ontario Public Health Convention (TOPHC), Toronto, ON, Canada. pp. 2–4.

Vos, T., Lim, S.S., Abbafati, C., Abbas, K.M., Abbasi, M., Abbasifard, M., Abbasi-Kangevari, M., Abbastabar, H., Abd-Allah, F., Abdelalim, A., Abdollahi, M., Abdollahpour, I., Abolhassani, H., Aboyans, V., Abrams, E.M., Abreu, L.G., Abrigo, M.R.M., Abu-Raddad, L.J., Abushouk, A.I., Acebedo, A., Ackerman, I.N., Adabi, M., Adamu, A.A., Adebayo, O.M., Adekanmbi, V., Adelson, J.D., Adetokunboh, O.O., Adham, D., Afshari, M., Afshin, A., Agardh, E.E., Agarwal, G., Agesa, K.M., Aghaali, M., Aghamir, S.M.K., Agrawal, A., Ahmad, T., Ahmadi, A., Ahmadi, M., Ahmadieh, H., Ahmadpour, E., Akalu, T.Y., Akinyemi, R.O., Akinyemiju, T., Akombi, B., Al-Aly, Z., Alam, K., Alam, N., Alam, S., Alam, T., Alanzi, T.M., Albertson, S.B., Alcalde-Rabanal, J.E., Alema, N.M., Ali, M., Ali, S., Alicandro, G., Alijanzadeh, M., Alinia, C., Alipour, V., Aljunid, S.M., Alla, F., Allebeck, P., Almasi-Hashiani, A., Alonso, J., Al-Raddadi, R.M., Altirkawi, K.A., Alvis-Guzman, N., Alvis-Zakzuk, N.J., Amini, S., Amini-Rarani, M., Aminorroaya, A., Amiri, F., Amit, A.M.L., Amugsi, D.A., Amul, G.G.H., Anderlini, D., Andrei, C.L., Andrei, T., Anjomshoa, M., Ansari, F., Ansari, I., Ansari-Moghaddam, A., Antonio, C.A.T., Antony, C.M., Antriyandarti, E., Anvari, D., Anwer, R., Arabloo, J., Arab-Zozani, M., Aravkin, A.Y., Ariani, F., Ärnlöv, J., Aryal, K.K., Arzani, A., Asadi-Aliabadi, M., Asadi-Pooya, A.A., Asghari, B., Ashbaugh, C., Atnafu, D.D., Atre, S.R., Ausloos, F., Ausloos, M., Ayala Quintanilla, B.P., Ayano, G., Ayanore, M.A., Aynalem, Y.A., Azari, S., Azarian, G., Azene, Z.N., Babaee, E., Badawi, A., Bagherzadeh, M., Bakhshaei, M.H., Bakhtiari, A., Balakrishnan, S., Balalla, S., Balassyano, S., Banach, M., Banik, P.C., Bannick, M.S., Bante, A.B., Baraki, A.G., Barboza, M.A., Barker-Collo, S.L., Barthelemy, C.M., Barua, L., Barzegar, A., Basu, S., Baune, B.T., Bayati, M., Bazmandegan, G., Bedi, N., Beghi, E., Béjot, Y., Bello, A.K., Bender, R.G., Bennett, D.A., Bennitt, F.B., Bensenor, I.M., Benziger, C.P., Berhe, K., Bernabe, E., Bertolacci, G.J., Bhageerathy, R., Bhala, N., Bhandari, D., Bhardwaj, P., Bhattacharyya, K., Bhutta, Z.A., Bibi, S., Biehl, M.H., Bikbov, B., Bin Sayeed, M.S., Biondi, A., Birihane, B.M., Bisanzio, D., Bisignano, C., Biswas, R.K., Bohlouli, S., Bohluli, M., Bolla, S.R.R., Boloor, A., Boon-Dooley, A.S., Borges, G., Borzì, A.M., Bourne, R., Brady, O.J., Brauer, M., Brayne, C., Breitborde, N.J.K., Brenner, H., Briant, P.S., Briggs, A.M., Briko, N.I., Britton, G.B., Bryazka, D., Buchbinder, R., Bumgarner, B.R., Busse, R., Butt, Z.A., Caetano dos Santos, F.L., Cámera, L.L.A.A., Campos-Nonato, I.R., Car, J., Cárdenas, R., Carreras, G., Carrero, J.J., Carvalho, F., Castaldelli-Maia, J.M., Castañeda-Orjuela, C.A., Castelpietra, G., Castle, C.D., Castro, F., Catalá-López, F., Causey, K., Cederroth, C.R., Cercy, K.M., Cerin, E., Chandan, J.S., Chang, A.R., Charlson, F.J., Chattu, V.K., Chaturvedi, S., Chimed-Ochir, O., Chin, K.L., Cho, D.Y., Christensen, H., Chu, D.-T., Chung, M.T., Cicuttini, F.M., Ciobanu, L.G., Cirillo, M., Collins, E.L., Compton, K., Conti, S., Cortesi, P.A., Costa, V.M., Cousin, E., Cowden, R.G., Cowie, B.C., Cromwell, E.A., Cross, D.H., Crowe, C.S., Cruz, J.A., Cunningham, M., Dahlawi, S.M.A., Damiani, G., Dandona, L., Dandona, R., Darwesh, A.M., Daryani, A., Das, J.K., Das Gupta, Rajat das Neves, J., Dávila-Cervantes, C.A., Davletov, K., De Leo, D., Dean, F.E., DeCleene, N.K., Deen, A., Degenhardt, L., Dellavalle, R.P., Demeke, F.M., Demsie, D.G., Denova-Gutiérrez, E., Dereje, N.D., Dervenis, N., Desai, R., Desalew, A., Dessie, G.A., Dharmaratne, S.D., Dhungana, G.P., Dianatinasab, M., Diaz, D., Dibaji Forooshani, Z.S., Dingels, Z. V, Dirac, M.A., Djalalinia, S., Do, H.T., Dokova, K., Dorostkar, F., Doshi, C.P., Doshmangir, L., Douiri, A., Doxey, M.C., Driscoll, T.R., Dunachie, S.J., Duncan, B.B., Duraes, A.R., Eagan, A.W., Ebrahimi Kalan, M., Edvardsson, D., Ehrlich, J.R., El Nahas, N., El Sayed, I., El Tantawi, M., Elbarazi, I., Elgendy, I.Y., Elhabashy, H.R., El-Jaafary, S.I., Elyazar, I.R.F., Emamian, M.H., Emmons-Bell, S., Erskine, H.E., Eshrati, B., Eskandarieh, S., Esmaeilnejad, S., Esmaeilzadeh, F., Esteghamati, A., Estep, K., Etemadi, A., Etisso, A.E., Farahmand, M., Faraj, A., Fareed, M., Faridnia, R., Farinha, C.S. e S., Farioli, A., Faro, A., Faruque, M., Farzadfar, F., Fattahi, N., Fazlzadeh, M., Feigin, V.L., Feldman, R., Fereshtehnejad, S.-M., Fernandes, E., Ferrari, A.J., Ferreira, M.L., Filip, I., Fischer, F., Fisher, J.L., Fitzgerald, R., Flohr, C., Flor, L.S., Foigt, N.A., Folayan, M.O., Force, L.M., Fornari, C., Foroutan, M., Fox, J.T., Freitas, M., Fu, W., Fukumoto, T., Furtado, J.M., Gad, M.M., Gakidou, E., Galles, N.C., Gallus, S., Gamkrelidze, A., Garcia-Basteiro, A.L., Gardner, W.M., Geberemariyam, B.S., Gebrehiwot, A.M., Gebremedhin, K.B., Gebreslassie, A.A.A.A., Gershberg Hayoon, A., Gething, P.W., Ghadimi, M., Ghadiri, K., Ghafourifard, M., Ghajar, A., Ghamari, F., Ghashghaee, A., Ghiasvand, H., Ghith, N., Gholamian, A., Gilani, S.A., Gill, P.S., Gitimoghaddam, M., Giussani, G., Goli, S., Gomez, R.S., Gopalani, S.V., Gorini, G., Gorman, T.M., Gottlich, H.C., Goudarzi, H., Goulart, A.C., Goulart, B.N.G., Grada, A., Grivna, M., Grosso, G., Gubari, M.I.M., Gugnani, H.C., Guimaraes, A.L.S., Guimarães, R.A., Guled, R.A., Guo, G., Guo, Y., Gupta, Rajeev, Haagsma, J.A., Haddock, B., Hafezi-Nejad, N., Hafiz, A., Hagins, H., Haile, L.M., Hall, B.J., Halvaei, I., Hamadeh, R.R., Hamagharib Abdullah, K., Hamilton, E.B., Han, C., Han, H., Hankey, G.J., Haro, J.M., Harvey, J.D., Hasaballah, A.I., Hasanzadeh, A., Hashemian, M., Hassanipour, S., Hassankhani, H., Havmoeller, R.J., Hay, R.J., Hay, S.I., Hayat, K., Heidari, B., Heidari, G., Heidari-Soureshjani, R., Hendrie, D., Henrikson, H.J., Henry, N.J., Herteliu, C., Heydarpour, F., Hird, T.R., Hoek, H.W., Hole, M.K., Holla, R., Hoogar, P., Hosgood, H.D., Hosseinzadeh, M., Hostiuc, M., Hostiuc, S., Househ, M., Hoy, D.G., Hsairi, M., Hsieh, V.C., Hu, G., Huda, T.M., Hugo, F.N., Huynh, C.K., Hwang, B.-F., Iannucci, V.C., Ibitoye, S.E., Ikuta, K.S., Ilesanmi, O.S., Ilic, I.M., Ilic, M.D., Inbaraj, L.R., Ippolito, H., Irvani, S.S.N., Islam, M.M., Islam, M., Islam, S.M.S., Islami, F., Iso, H., Ivers, R.Q., Iwu, C.C.D., Iyamu, I.O., Jaafari, J., Jacobsen, K.H., Jadidi-Niaragh, F., Jafari, H., Jafarinia, M., Jahagirdar, D., Jahani, M.A., Jahanmehr, N., Jakovljevic, M., Jalali, A., Jalilian, F., James, S.L., Janjani, H., Janodia, M.D., Jayatilleke, A.U., Jeemon, P., Jenabi, E., Jha, R.P., Jha, V., Ji, J.S., Jia, P., John, O., John-Akinola, Y.O., Johnson, C.O., Johnson, S.C., Jonas, J.B., Joo, T., Joshi, A., Jozwiak, J.J., Jürisson, M., Kabir, A., Kabir, Z., Kalani, H., Kalani, R., Kalankesh, L.R., Kalhor, R., Kamiab, Z., Kanchan, T., Karami Matin, B., Karch, A., Karim, M.A., Karimi, S.E., Kassa, G.M., Kassebaum, N.J., Katikireddi, S.V., Kawakami, N., Kayode, G.A., Keddie, S.H., Keller, C., Kereselidze, M., Khafaie, M.A., Khalid, N., Khan, M., Khatab, K., Khater, M.M., Khatib, M.N., Khayamzadeh, M., Khodayari, M.T., Khundkar, R., Kianipour, N., Kieling, C., Kim, D., Kim, Y.-E., Kim, Y.J., Kimokoti, R.W., Kisa, A., Kisa, S., Kissimova-Skarbek, K., Kivimäki, M., Kneib, C.J., Knudsen, A.K.S., Kocarnik, J.M., Kolola, T., Kopec, J.A., Kosen, S., Koul, P.A., Koyanagi, A., Kravchenko, M.A., Krishan, K., Krohn, K.J., Kuate Defo, B., Kucuk Bicer, B., Kumar, G.A., Kumar, M., Kumar, P., Kumar, V., Kumaresh, G., Kurmi, O.P., Kusuma, D., Kyu, H.H., La Vecchia, C., Lacey, B., Lal, D.K., Lalloo, R., Lam, J.O., Lami, F.H., Landires, I., Lang, J.J., Lansingh, V.C., Larson, S.L., Larsson, A.O., Lasrado, S., Lassi, Z.S., Lau, K.M.-M., Lavados, P.M., Lazarus, J. V, Ledesma, J.R., Lee, P.H., Lee, S.W.H., LeGrand, K.E., Leigh, J., Leonardi, M., Lescinsky, H., Leung, J., Levi, M., Lewington, S., Li, S., Lim, L.-L., Lin, C., Lin, R.-T., Linehan, C., Linn, S., Liu, H.-C., Liu, S., Liu, Z., Looker, K.J., Lopez, A.D., Lopukhov, P.D., Lorkowski, S., Lotufo, P.A., Lucas, T.C.D., Lugo, A., Lunevicius, R., Lyons, R.A., Ma, J., MacLachlan, J.H., Maddison, E.R., Maddison, R., Madotto, F., Mahasha, P.W., Mai, H.T., Majeed, A., Maled, V., Maleki, S., Malekzadeh, R., Malta, D.C., Mamun, A.A., Manafi, A., Manafi, N., Manguerra, H., Mansouri, B., Mansournia, M.A., Mantilla Herrera, A.M., Maravilla, J.C., Marks, A., Martins-Melo, F.R., Martopullo, I., Masoumi, S.Z., Massano, J., Massenburg, B.B., Mathur, M.R., Maulik, P.K., McAlinden, C., McGrath, J.J., McKee, M., Mehndiratta, M.M., Mehri, F., Mehta, K.M., Meitei, W.B., Memiah, P.T.N., Mendoza, W., Menezes, R.G., Mengesha, E.W., Mengesha, M.B., Mereke, A., Meretoja, A., Meretoja, T.J., Mestrovic, T., Miazgowski, B., Miazgowski, T., Michalek, I.M., Mihretie, K.M., Miller, T.R., Mills, E.J., Mirica, A., Mirrakhimov, E.M., Mirzaei, H., Mirzaei, M., Mirzaei-Alavijeh, M., Misganaw, A.T., Mithra, P., Moazen, B., Moghadaszadeh, M., Mohamadi, E., Mohammad, D.K., Mohammad, Y., Mohammad Gholi Mezerji, N., Mohammadian-Hafshejani, A., Mohammadifard, N., Mohammadpourhodki, R., Mohammed, S., Mokdad, A.H., Molokhia, M., Momen, N.C., Monasta, L., Mondello, S., Mooney, M.D., Moosazadeh, M., Moradi, G., Moradi, M., Moradi-Lakeh, M., Moradzadeh, R., Moraga, P., Morales, L., Morawska, L., Moreno Velásquez, I., Morgado-da-Costa, J., Morrison, S.D., Mosser, J.F., Mouodi, S., Mousavi, S.M., Mousavi Khaneghah, A., Mueller, U.O., Munro, S.B., Muriithi, M.K., Musa, K.I., Muthupandian, S., Naderi, M., Nagarajan, A.J., Nagel, G., Naghshtabrizi, B., Nair, S., Nandi, A.K., Nangia, V., Nansseu, J.R., Nayak, V.C., Nazari, J., Negoi, I., Negoi, R.I., Netsere, H.B.N., Ngunjiri, J.W., Nguyen, C.T., Nguyen, J., Nguyen, Michele, Nguyen, Minh, Nichols, E., Nigatu, D., Nigatu, Y.T., Nikbakhsh, R., Nixon, M.R., Nnaji, C.A., Nomura, S., Norrving, B., Noubiap, J.J., Nowak, C., Nunez-Samudio, V., Otoiu, A., Oancea, B., Odell, C.M., Ogbo, F.A., Oh, I.-H., Okunga, E.W., Oladnabi, M., Olagunju, A.T., Olusanya, B.O., Olusanya, J.O., Oluwasanu, M.M., Omar Bali, A., Omer, M.O., Ong, K.L., Onwujekwe, O.E., Orji, A.U., Orpana, H.M., Ortiz, A., Ostroff, S.M., Otstavnov, N., Otstavnov, S.S., Øverland, S., Owolabi, M.O. P A M., Padubidri, J.R., Pakhare, A.P., Palladino, R., Pana, A., Panda-Jonas, S., Pandey, A., Park, E.-K., Parmar, P.G.K., Pasupula, D.K., Patel, S.K., Paternina-Caicedo, A.J., Pathak, A., Pathak, M., Patten, S.B., Patton, G.C., Paudel, D., Pazoki Toroudi, H., Peden, A.E., Pennini, A., Pepito, V.C.F., Peprah, E.K., Pereira, A., Pereira, D.M., Perico, N., Pham, H.Q., Phillips, M.R., Pigott, D.M., Pilgrim, T., Pilz, T.M., Pirsaheb, M., Plana-Ripoll, O., Plass, D., Pokhrel, K.N., Polibin, R. V, Polinder, S., Polkinghorne, K.R., Postma, M.J., Pourjafar, H., Pourmalek, F., Pourmirza Kalhori, R., Pourshams, A., Poznańska, A., Prada, S.I., Prakash, V., Pribadi, D.R.A., Pupillo, E., Quazi Syed, Z., Rabiee, M., Rabiee, N., Radfar, A., Rafiee, A., Rafiei, A., Raggi, A., Rahimi-Movaghar, A., Rahman, M.A., Rajabpour-Sanati, A., Rajati, F., Ramezanzadeh, K., Ranabhat, C.L., Rao, P.C., Rao, S.J., Rasella, D., Rastogi, P., Rathi, P., Rawaf, D.L., Rawaf, S., Rawal, L., Razo, C., Redford, S.B., Reiner Jr, R.C., Reinig, N., Reitsma, M.B., Remuzzi, G., Renjith, V., Renzaho, A.M.N., Resnikoff, S., Rezaei, N., Rezai, M. sadegh, Rezapour, A., Rhinehart, P.-A., Riahi, S.M., Ribeiro, A.L.P., Ribeiro, D.C., Ribeiro, D., Rickard, J., Roberts, N.L.S., Roberts, S., Robinson, S.R., Roever, L., Rolfe, S., Ronfani, L., Roshandel, G., Roth, G.A., Rubagotti, E., Rumisha, S.F., Sabour, S., Sachdev, P.S., Saddik, B., Sadeghi, E., Sadeghi, M., Saeidi, S., Safi, S., Safiri, S., Sagar, R., Sahebkar, A., Sahraian, M.A., Sajadi, S.M., Salahshoor, M.R., Salamati, P., Salehi Zahabi, S., Salem, H., Salem, M.R.R., Salimzadeh, H., Salomon, J.A., Salz, I., Samad, Z., Samy, A.M., Sanabria, J., Santomauro, D.F., Santos, I.S., Santos, J.V., Santric-Milicevic, M.M., Saraswathy, S.Y.I., Sarmiento-Suárez, R., Sarrafzadegan, N., Sartorius, B., Sarveazad, A., Sathian, B., Sathish, T., Sattin, D., Sbarra, A.N., Schaeffer, L.E., Schiavolin, S., Schmidt, M.I., Schutte, A.E., Schwebel, D.C., Schwendicke, F., Senbeta, A.M., Senthilkumaran, S., Sepanlou, S.G., Shackelford, K.A., Shadid, J., Shahabi, S., Shaheen, A.A., Shaikh, M.A., Shalash, A.S., Shams-Beyranvand, M., Shamsizadeh, M., Shannawaz, M., Sharafi, K., Sharara, F., Sheena, B.S., Sheikhtaheri, A., Shetty, R.S., Shibuya, K., Shiferaw, W.S., Shigematsu, M., Shin, J. Il, Shiri, R., Shirkoohi, R., Shrime, M.G., Shuval, K., Siabani, S., Sigfusdottir, I.D., Sigurvinsdottir, R., Silva, J.P., Simpson, K.E., Singh, A., Singh, J.A., Skiadaresi, E., Skou, S.T.S., Skryabin, V.Y., Sobngwi, E., Sokhan, A., Soltani, S., Sorensen, R.J.D., Soriano, J.B., Sorrie, M.B., Soyiri, I.N., Sreeramareddy, C.T., Stanaway, J.D., Stark, B.A., Stefan, S.C., Stein, C., Steiner, C., Steiner, T.J., Stokes, M.A., Stovner, L.J., Stubbs, J.L., Sudaryanto, A., Sufiyan, M.B., Sulo, G., Sultan, I., Sykes, B.L., Sylte, D.O., Szócska, M., Tabarés-Seisdedos, R., Tabb, K.M., Tadakamadla, S.K., Taherkhani, A., Tajdini, M., Takahashi, K., Taveira, N., Teagle, W.L., Teame, H., Tehrani-Banihashemi, A., Teklehaimanot, B.F., Terrason, S., Tessema, Z.T., Thankappan, K.R., Thomson, A.M., Tohidinik, H.R., Tonelli, M., Topor-Madry, R., Torre, A.E., Touvier, M., Tovani-Palone, M.R.R., Tran, B.X., Travillian, R., Troeger, C.E., Truelsen, T.C., Tsai, A.C., Tsatsakis, A., Tudor Car, L., Tyrovolas, S., Uddin, R., Ullah, S., Undurraga, E.A., Unnikrishnan, B., Vacante, M., Vakilian, A., Valdez, P.R., Varughese, S., Vasankari, T.J., Vasseghian, Y., Venketasubramanian, N., Violante, F.S., Vlassov, V., Vollset, S.E., Vongpradith, A., Vukovic, A., Vukovic, R., Waheed, Y., Walters, M.K., Wang, J., Wang, Y., Wang, Y.-P., Ward, J.L., Watson, A., Wei, J., Weintraub, R.G., Weiss, D.J., Weiss, J., Westerman, R., Whisnant, J.L., Whiteford, H.A., Wiangkham, T., Wiens, K.E., Wijeratne, T., Wilner, L.B., Wilson, S., Wojtyniak, B., Wolfe, C.D.A., Wool, E.E., Wu, A.-M., Wulf Hanson, S., Wunrow, H.Y., Xu, G., Xu, R., Yadgir, S., Yahyazadeh Jabbari, S.H., Yamagishi, K., Yaminfirooz, M., Yano, Y., Yaya, S., Yazdi-Feyzabadi, V., Yearwood, J.A., Yeheyis, T.Y., Yeshitila, Y.G., Yip, P., Yonemoto, N., Yoon, S.-J., Yoosefi Lebni, J., Younis, M.Z., Younker, T.P., Yousefi, Z., Yousefifard, M., Yousefinezhadi, T., Yousuf, A.Y., Yu, C., Yusefzadeh, H., Zahirian Moghadam, T., Zaki, L., Zaman, S. Bin, Zamani, M., Zamanian, M., Zandian, H., Zangeneh, A., Zastrozhin, M.S., Zewdie, K.A., Zhang, Y., Zhang, Z.-J., Zhao, J.T., Zhao, Y., Zheng, P., Zhou, M., Ziapour, A., Zimsen, S.R.M., Naghavi, M., Murray, C.J.L., 2020. Global burden of 369 diseases and injuries in 204 countries and territories, 1990–2019: a systematic analysis for the Global Burden of Disease Study 2019. Lancet 396, 1204–1222. https://doi.org/10.1016/S0140-6736(20)30925-9

World Health Organisation, 2017. Guidelines for drinking-water quality, 4th edition: 1st addendum, World Health Organization. WHO.

