## Supplementary Material for "Burden of Disease from Contaminated Drinking Water in Countries with High Access to Safely Managed Water: A Systematic Review"

**Supplementary Table S1**. Countries and territories with ≥90% of population using safely managed drinking water services

| Country/Territory | People using safely managed drinking water services (% of population in 2017)[1] |
|---|---|
| Andorra | 90.6 |
| Austria | 98.9 |
| Belgium | 99.5 |
| Bulgaria | 96.9 |
| Bahrain | 99.0 |
| Belarus | 94.5 |
| Canada | 98.9 |
| Channel Islands | 92.0 |
| Chile | 98.6 |
| Costa Rica | 93.8 |
| Cyprus | 99.6 |
| Czech Republic | 97.9 |
| Germany | 99.8 |
| Denmark | 96.7 |
| Estonia | 93.3 |
| Finland | 99.6 |
| France | 97.9 |
| United Kingdom | 100.0 |
| Gibraltar | 100.0 |
| Greece | 100.0 |
| Greenland | 96.7 |
| Guam | 99.5 |
| Hong Kong SAR, China | 100.0 |
| Isle of Man | 97.2 |
| Ireland | 97.3 |
| Iran, Islamic Rep. | 91.8 |
| Iceland | 100.0 |
| Israel | 99.4 |
| Italy | 95.0 |
| Jordan | 93.8 |
| Japan | 98.5 |
| Korea, Rep. | 98.2 |

[1]People using safely managed drinking water services (% of population) refers to the percentage of people using drinking water from an improved source that is accessible on premises, available when needed and free from fecal and priority chemical contamination. Improved water sources include piped water, boreholes or tube wells, protected dug wells, protected springs, and packaged or delivered water, according to the WHO/UNICEF Joint Monitoring Program (JMP) for Water Supply, Sanitation and Hygiene.

| | |
|---|---|
| Kuwait | 100.0 |
| Liechtenstein | 100.0 |
| Lithuania | 92.0 |
| Luxembourg | 99.7 |
| Latvia | 95.2 |
| Macao SAR, China | 100.0 |
| Monaco | 100.0 |
| Malta | 100.0 |
| Montenegro | 93.6 |
| Northern Mariana Islands | 90.2 |
| Malaysia | 93.3 |
| New Caledonia | 96.7 |
| Netherlands | 100.0 |
| Norway | 98.3 |
| New Zealand | 100.0 |
| Oman | 90.3 |
| Poland | 99.2 |
| Puerto Rico | 94.1 |
| Portugal | 95.3 |
| Qatar | 96.2 |
| Singapore | 100.0 |
| San Marino | 100.0 |
| Slovak Republic | 99.8 |
| Slovenia | 98.1 |
| Sweden | 99.9 |
| Switzerland | 95.5 |
| Spain | 98.4 |
| Turkmenistan | 93.9 |
| Tunisia | 92.7 |
| Ukraine | 92.0 |
| United States | 99.0 |

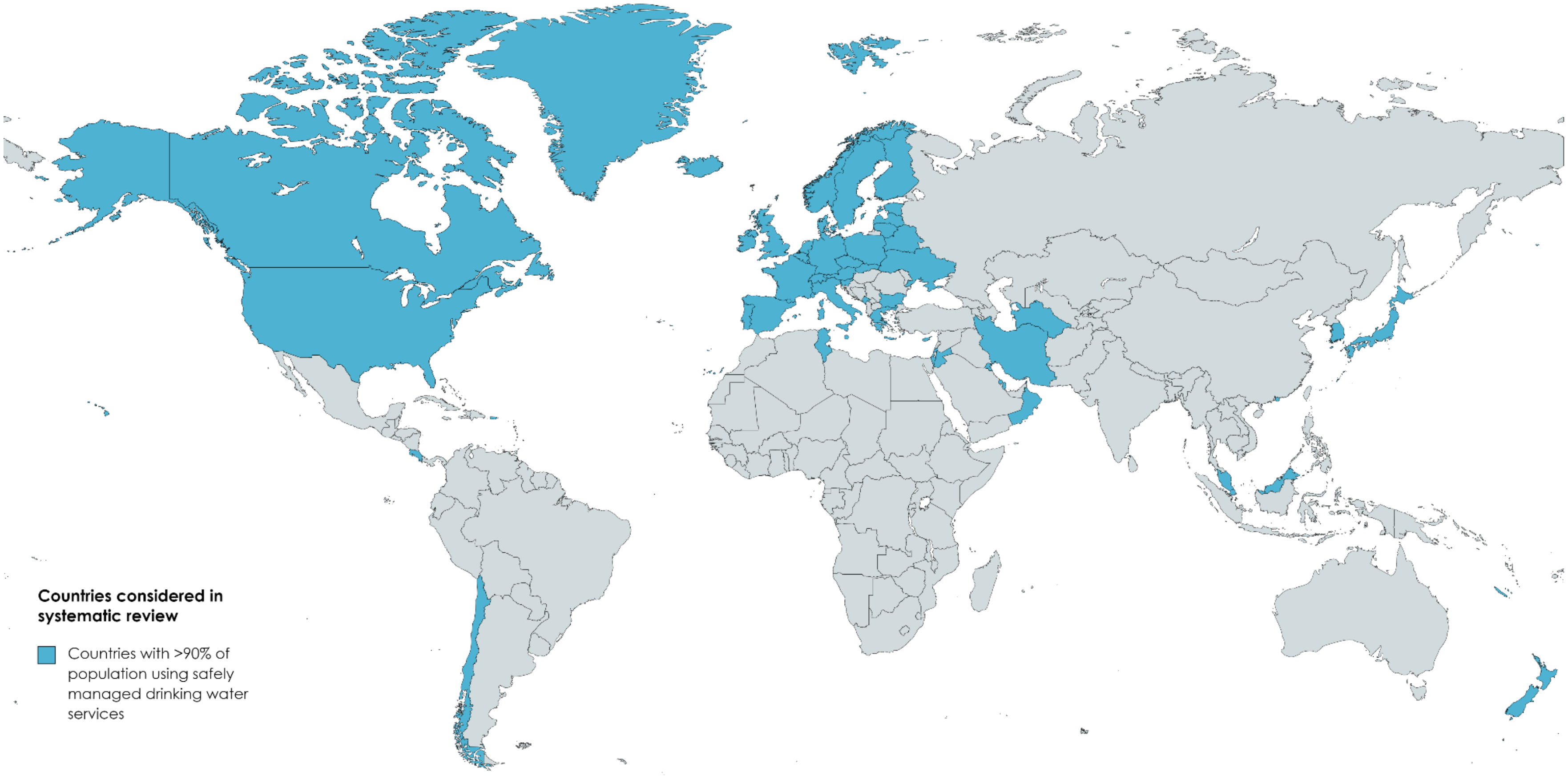


**Supplementary Figure S1**. Countries with ≥90% of population using safely managed drinking water service

**Supplementary Table S2.** PRISMA Checklist

| Section and Topic | Item # | Checklist item | Location where item is reported |
|---|---|---|---|
| **TITLE** | | | |
| Title | 1 | Identify the report as a systematic review. | p. 1 |
| **ABSTRACT** | | | |
| Abstract | 2 | See the PRISMA 2020 for Abstracts checklist. | Items 1-4, 6-10, 11 in abstract; all other items are NA |
| **INTRODUCTION** | | | |
| Rationale | 3 | Describe the rationale for the review in the context of existing knowledge. | pp. 2-3 (first four paragraphs of “Introduction”) |
| Objectives | 4 | Provide an explicit statement of the objective(s) or question(s) the review addresses. | p. 3 (last paragraph of “Introduction)” |
| **METHODS** | | | |
| Eligibility criteria | 5 | Specify the inclusion and exclusion criteria for the review and how studies were grouped for the syntheses. | p. 3 (Section 2.2 and 2.3) |
| Information sources | 6 | Specify all databases, registers, websites, organisations, reference lists and other sources searched or consulted to identify studies. Specify the date when each source was last searched or consulted. | p. 3 (Section 2.2) |
| Search strategy | 7 | Present the full search strategies for all databases, registers and websites, including any filters and limits used. | p. 3 (Section 2.2) |
| Selection process | 8 | Specify the methods used to decide whether a study met the inclusion criteria of the review, including how many reviewers screened each record and each report retrieved, whether they worked independently, and if applicable, details of automation tools used in the process. | p. 3 (Section 2.3) |
| Data collection process | 9 | Specify the methods used to collect data from reports, including how many reviewers collected data from each report, whether they worked independently, any processes for obtaining or confirming data from study investigators, and if applicable, details of automation tools used in the process. | pp. 3-4 (Section 2.4) |
| Data items | 10a | List and define all outcomes for which data were sought. Specify whether all results that were compatible with each outcome domain in each study were sought (e.g. for all measures, time points, analyses), and if not, the methods used to decide which results to collect. | pp. 3-4 (Section 2.4) |
| | 10b | List and define all other variables for which data were sought (e.g. participant and intervention characteristics, funding sources). Describe any assumptions made about any missing or unclear information. | pp. 3-4 (Section 2.4) |
| Study risk of bias assessment | 11 | Specify the methods used to assess risk of bias in the included studies, including details of the tool(s) used, how many reviewers assessed each study and whether they worked independently, and if applicable, details of automation tools used in the process. | NA |

| Section and Topic | Item # | Checklist item | Location where item is reported |
|---|---|---|---|
| Effect measures | 12 | Specify for each outcome the effect measure(s) (e.g. risk ratio, mean difference) used in the synthesis or presentation of results. | pp. 3-4 (Section 2.4) |
| Synthesis methods | 13a | Describe the processes used to decide which studies were eligible for each synthesis (e.g. tabulating the study intervention characteristics and comparing against the planned groups for each synthesis (item #5)). | pp. 3-4 (Section 2.4) |
| | 13b | Describe any methods required to prepare the data for presentation or synthesis, such as handling of missing summary statistics, or data conversions. | pp. 3-4 (Section 2.4) |
| | 13c | Describe any methods used to tabulate or visually display results of individual studies and syntheses. | Supplementary Text S2 |
| | 13d | Describe any methods used to synthesize results and provide a rationale for the choice(s). If meta-analysis was performed, describe the model(s), method(s) to identify the presence and extent of statistical heterogeneity, and software package(s) used. | pp. 3-4 (Section 2.4) and Supplementary Text S2 |
| | 13e | Describe any methods used to explore possible causes of heterogeneity among study results (e.g. subgroup analysis, meta-regression). | NA |
| | 13f | Describe any sensitivity analyses conducted to assess robustness of the synthesized results. | NA |
| Reporting bias assessment | 14 | Describe any methods used to assess risk of bias due to missing results in a synthesis (arising from reporting biases). | NA |
| Certainty assessment | 15 | Describe any methods used to assess certainty (or confidence) in the body of evidence for an outcome. | NA |
| **RESULTS** | | | |
| Study selection | 16a | Describe the results of the search and selection process, from the number of records identified in the search to the number of studies included in the review, ideally using a flow diagram. | p. 4 (Section 3) Supplementary Figure S2 |
| | 16b | Cite studies that might appear to meet the inclusion criteria, but which were excluded, and explain why they were excluded. | NA |
| Study characteristics | 17 | Cite each included study and present its characteristics. | Tables 1 and 2 |
| Risk of bias in studies | 18 | Present assessments of risk of bias for each included study. | NA |
| Results of individual studies | 19 | For all outcomes, present, for each study: (a) summary statistics for each group (where appropriate) and (b) an effect estimate and its precision (e.g. confidence/credible interval), ideally using structured tables or plots. | Figures 1 and 2 |
| Results of syntheses | 20a | For each synthesis, briefly summarise the characteristics and risk of bias among contributing studies. | NA |
| | 20b | Present results of all statistical syntheses conducted. If meta-analysis was done, present for each the summary estimate and its precision (e.g. confidence/credible interval) and measures of statistical heterogeneity. If comparing groups, describe the direction of the effect. | pp. 4-5 (Sections 3.1 and 3.2) |

| Section and Topic | Item # | Checklist item | Location where item is reported |
|---|---|---|---|
| | 20c | Present results of all investigations of possible causes of heterogeneity among study results. | NA |
| | 20d | Present results of all sensitivity analyses conducted to assess the robustness of the synthesized results. | NA |
| Reporting biases | 21 | Present assessments of risk of bias due to missing results (arising from reporting biases) for each synthesis assessed. | NA |
| Certainty of evidence | 22 | Present assessments of certainty (or confidence) in the body of evidence for each outcome assessed. | NA |
| **DISCUSSION** | | | |
| Discussion | 23a | Provide a general interpretation of the results in the context of other evidence. | pp. 5-11 |
| | 23b | Discuss any limitations of the evidence included in the review. | pp. 7-10 (Section 4.1) |
| | 23c | Discuss any limitations of the review processes used. | pp. 10-11 (Section 4.1.5) |
| | 23d | Discuss implications of the results for practice, policy, and future research. | p. 11 (Section 4.1.5) |
| **OTHER INFORMATION** | | | |
| Registration and protocol | 24a | Provide registration information for the review, including register name and registration number, or state that the review was not registered. | NA |
| | 24b | Indicate where the review protocol can be accessed, or state that a protocol was not prepared. | p. 12 |
| | 24c | Describe and explain any amendments to information provided at registration or in the protocol. | NA |
| Support | 25 | Describe sources of financial or non-financial support for the review, and the role of the funders or sponsors in the review. | pp 11-12 |
| Competing interests | 26 | Declare any competing interests of review authors. | p. 12 |
| Availability of data, code and other materials | 27 | Report which of the following are publicly available and where they can be found: template data collection forms; data extracted from included studies; data used for all analyses; analytic code; any other materials used in the review. | p. 11 |

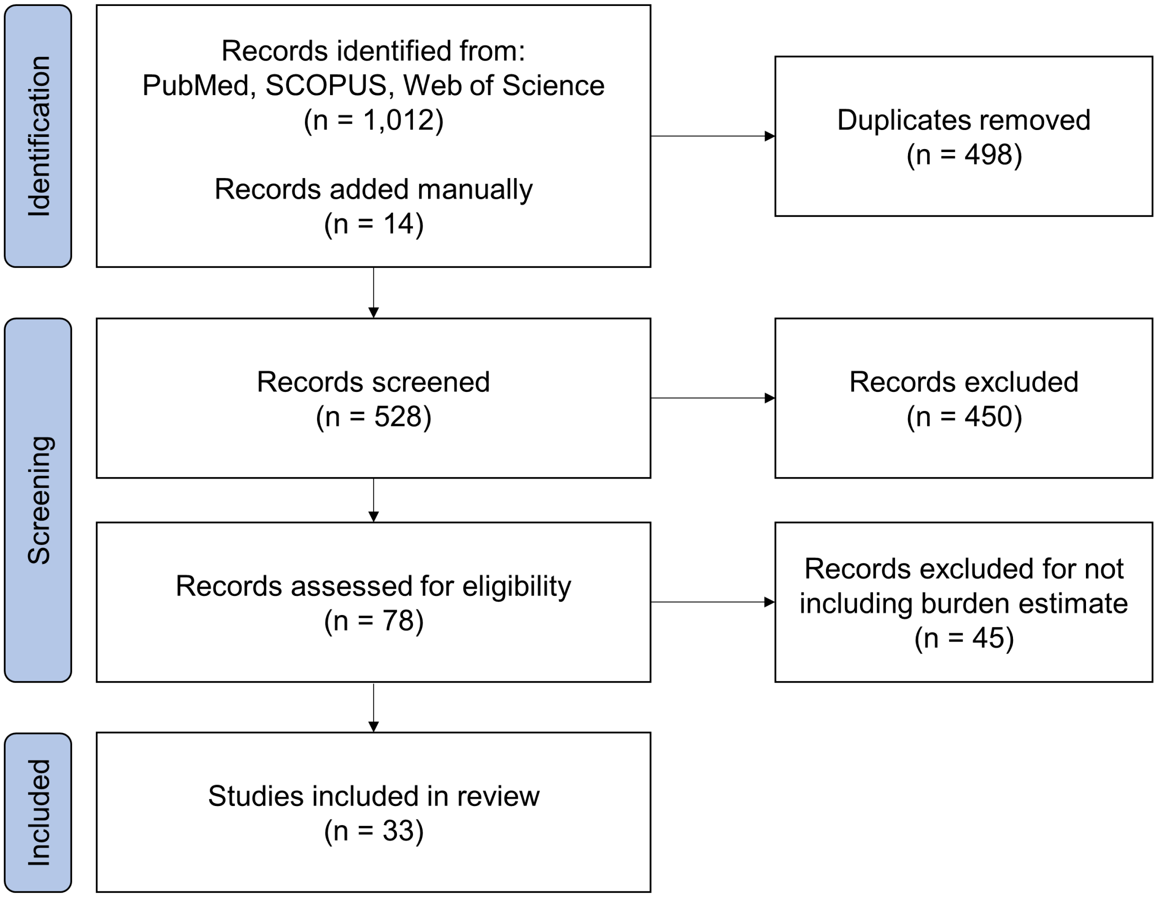


**Supplementary Figure S2**. Flow chart of study selection for review

**Supplementary Text S1.** A description of the development of major concepts, countries, and search terms for the systematic review.

**Initial Search in PubMed:**

**#1. Concept A:** “Burden of disease”

**#2. Concept B:** “drinking water” OR “tap water”

**#3. Concept 3:** Countries with proportion of population using safely managed drinking water services ≥75% (2017)

“developed country” OR “developed countries” OR “Europe” OR “UK” OR “USA” OR “United States” OR “Australia” OR “United Kingdom” OR "Malta" OR "Gibraltar" OR "Greece" OR "Hong Kong" OR "Kuwait" OR "Liechtenstein" OR "Macao" OR "Monaco" OR "New Zealand" OR "Singapore" OR "San Marino" OR "Iceland" OR "Great Britain and Northern Ireland" OR "Netherlands" OR "Sweden" OR "Germany" OR "Slovakia" OR "Luxembourg" OR "Finland" OR "Cyprus" OR "Guam" OR "Belgium" OR "Israel" OR "Poland" OR "United States of America" OR "Bahrain" OR "Austria" OR "Martinique" OR "Canada" OR "Chile" OR "Japan" OR "Spain" OR "Norway" OR "Republic of Korea" OR "Slovenia" OR "Czechia" OR "France" OR "Guadeloupe" OR "Ireland" OR "Niue" OR "Isle of Man" OR "Bulgaria" OR "Greenland" OR "Denmark" OR "New Caledonia" OR "Qatar" OR " Réunion" OR "Switzerland" OR "Portugal" OR "Latvia" OR "Italy" OR "Belarus" OR "Puerto Rico" OR "Turkmenistan" OR "Jordan" OR "Costa Rica" OR "Montenegro" OR "Estonia" OR "Malaysia" OR "Tunisia" OR "Lithuania" OR "Ukraine" OR "Channel Islands" OR "Iran" OR "French Guiana" OR "Andorra" OR "Oman" OR "Northern Mariana Islands" OR "Croatia" OR "Hungary" OR "Kazakhstan" OR "Saint Helena" OR "Bosnia and Herzegovina" OR "Grenada" OR "Armenia" OR "Mayotte" OR "Saint Pierre and Miquelon" OR "Romania" OR "North Macedonia" OR "Georgia" OR "Russian Federation" OR "Ecuador"

**#4. Combine the three concepts: #1 AND #2 AND #3**

| Searches | Result (i.e., # of articles) |
|---|---|
| #1 (see above) | 11,418 |
| #2 (see above) | 62,419 |
| #3 (see above) | 16,970,406 |
| #4. Combined | **54** |

**Final Search in PubMed:**

**#1. Concept A:** "burden of disease" OR "disease burden" OR "gastrointestinal illness"

**#2. Concept B:** “drinking water” OR “tap water”

**#3. Concept 3:** Countries with proportion of population using safely managed drinking water services ≥90% (2017)

“developed country” OR “developed countries” OR “Europe” OR “UK” OR “USA” OR “United States” OR “Australia” OR “United Kingdom” OR "Malta" OR "Gibraltar" OR "Greece" OR "Hong Kong" OR "Kuwait" OR "Liechtenstein" OR "Macao" OR "Monaco" OR "New Zealand" OR "Singapore" OR "San Marino" OR "Iceland" OR "Great Britain and Northern Ireland" OR "Netherlands" OR "Sweden" OR "Germany" OR "Slovakia" OR "Luxembourg" OR "Finland" OR "Cyprus" OR "Guam" OR "Belgium" OR "Israel" OR "Poland" OR "United States of America" OR "Bahrain" OR "Austria" OR "Martinique" OR "Canada" OR "Chile" OR "Japan" OR "Spain" OR "Norway" OR "Republic of Korea" OR "Slovenia" OR "Czechia" OR "France" OR "Guadeloupe" OR "Ireland" OR "Niue" OR "Isle of Man" OR "Bulgaria" OR "Greenland" OR "Denmark" OR "New Caledonia" OR "Qatar" OR " Réunion" OR "Switzerland" OR "Portugal" OR "Latvia" OR "Italy" OR "Belarus" OR "Puerto Rico" OR "Turkmenistan" OR "Jordan" OR "Costa Rica" OR "Montenegro" OR "Estonia" OR "Malaysia" OR "Tunisia" OR "Lithuania" OR "Ukraine" OR "Channel Islands" OR "Iran" OR "French Guiana" OR "Andorra" OR "Oman" OR "Northern Mariana Islands"

**#4. Combine the three concepts: #1 AND #2 AND #3**

| **Searches** | **Result (i.e., # of articles)** |
|---|---|
| **#1 (see above)** | 27,433 |
| **#2 (see above)** | 62,444 |
| **#3 (see above)** | 16,812,709 |
| **#4. Combined** | **218** |

**Supplementary Table S3.** Explanations of scale factors used to estimate illness cases and deaths.

| Author | Applicable Population | Health Outcome Measure | Estimated Illness Cases | Estimated Deaths | Scale Factor | Annual Cases Per 100,000 | Annual Deaths Per 100,000 | Computation of Scale Factor |
|---|---|---|---|---|---|---|---|---|
| Beaudeau et al. 2014 | 201,000 | AGI hospital admissions (age >64 years) | 153 | NA | 50.0 | 3,806 | NA | 2% of U.S. AGI cases resulted in hospitalization[1] |
| Collier et al. 2020 | 318,600,000 | Cases of 8 waterborne GI illnesses (campylobacteriosis, cryptosporidiosis, giardiasis, norovirus, STEC infections, shigellosis, *Vibrio* infections) | 2,381,660 | 194 | 0.12 | 89.70 | 0.007 | The study reports the burden attributed to specific illnesses; summed total GI only; estimate was then scaled to reflect that 12% of GI can be attributed to drinking water (attributable risk estimated by Payment et al. 1997 and Colford et al. 2006) |
| DeFelice et al. 2016 | 9,459,000 | Emergency department visits for AGI | 29,400 | NA | 14.7 | 4,573 | NA | 405,000 emergency department visits for AGI were reported in NC per year during this study. From national AGI incidence rate of 0.63 cases/person-year, would expect 0.63*9.459 million = 5.959 million cases per year. 14.7 = $(5.959x10^6)$/ 405,000. |
| DeFelice et al., 2015, methods 1&2 | 7,516,380 | Emergency department visits for AGI | 190 | NA | 14.7 | 37.19 | NA | See DeFelice et al., 2016 |

| | | | | | | | | |
|---|---|---|---|---|---|---|---|---|
| DeFelice et al., 2015, method 3 | 7,516,380 | Emergency department visits for AGI | 32,000 | NA | 14.7 | 6,264 | NA | See DeFelice et al., 2016 |
| Gibney et al. 2017 | 22,300,000 | Cases of five waterborne GI illnesses | 7,179,590 | 10 | 0.19 | 6,117 | 0.01 | The study reports the burden from drinking water and recreational water; it cites information that 19% of reported waterborne GI outbreaks are from drinking water |
| MacDonald Gibson et al. 2013 | 4,448,000 | Cases of bladder and colorectal cancers | Bladder: 154 Colorectal: 328 | Bladder: 3 Colorectal: 9 | Bladder: 0.08 Colorectal: 0.06 | 0.75 | 0.27 | Scaled to cases using visits per patient data from Yabroff et al.[2] (11.8 visits per bladder cancer case; 16.1 visits per colorectal cancer case) |
| MacDonald Gibson et al. 2013 | 4,448,000 | GI medical facility visits | 46,200 | 4 | 2.04 | 2,120 | $9.0 \times 10^{-2}$ | 2017 study in UAE by Al Alkeem et al.[3] found 49% of those with infectious intestinal illnesses seek medical attention; 1/0.49=2.04. |
| Masciopinto et al. 2019 | 1,786,446 | GI illnesses arising from adenovirus, norovirus, enterovirus, and rotavirus | 348 | NA | 16.2 | 315.6 | NA | Published estimate is for waterborne virus exposure in summer (45 days/year). Scaled by 365/45 to reflect full year and by 1/0.50 to reflect average data from across studies in this review on fraction of waterborne cases attributable to viruses. For each study in which burden was |

| | | | | | | | | |
|---|---|---|---|---|---|---|---|---|
| | | | | | | | | estimated for specific etiologic agents and viral agents were included, a viral fraction was estimated and then averaged across studies. These studies included: Burch et al. 2021, Collier et al. 2021, Eisenberg et al. 2006, Gibney et al. 2017, Moore et al. 2010, Morris and Levin 1995, Murphy et al. 2016, and Verhougstraete et al. 2020 |

References


1 Imhoff B, Morse D, Shiferaw B, et al. Burden of Self-Reported Acute Diarrheal Illness in FoodNet Surveillance Areas, 1998–1999. *Clin Infect Dis* 2004; **38**: S219–26.

2 Yabroff KR, Davis WW, Lamont EB, et al. Patient Time Costs Associated With Cancer Care. *JNCI J Natl Cancer Inst* 2007; **99**: 14–23.

3 Al Alkeem F, Loney T, Aziz F, Blair I, Sonnevend Á, Sheek-Hussein M. Prevalence and factors associated with infectious intestinal diseases in Ras Al Khaimah, United Arab Emirates, 2017: A population-based cross-sectional study. *Int J Infect Dis* 2019; **85**: 188–94.

**Supplementary Text S2. Data Synthesis and Visualization**

Burden estimates by country/territory were plotted for burden of disease from microbiological and chemical contamination of drinking water. Confidence intervals were not available for these estimates. Plots were generated to demonstrate the extent of estimates available by country, contaminant, and disease outcome. Population-weighted average disease burden was estimated for disease outcomes.

**Supplementary Table S4**. Global distribution of studies included in final review.

| Country | Number of studies |
|---|---|
| United States | 18 |
| Canada | 5 |
| New Zealand | 3 |
| Iran | 2 |
| Australia | 1 |
| European Union (EU28[2]) | 1 |
| Italy | 1 |
| Korea | 1 |
| Netherlands | 1 |
| United Kingdom | 1 |
| United Arab Emirates | 1 |

[2] EU28 refers to the 28 countries that were part of the European Union as of 2013, including Austria, Belgium, Bulgaria, Croatia, Cyprus, Czechia, Denmark, Estonia, Finland, France, Germany, Greece, Hungary, Ireland, Italy, Latvia, Lithuania, Luxembourg, Malta, Netherlands, Poland, Portugal, Romania, Slovakia, Slovenia, Spain, Sweden, and the United Kingdom (removed Feb. 2020). Of those, the following countries do not meet the ≥90% of population using safely managed drinking water services definition considered in our review: Croatia, Hungary, and Romania.

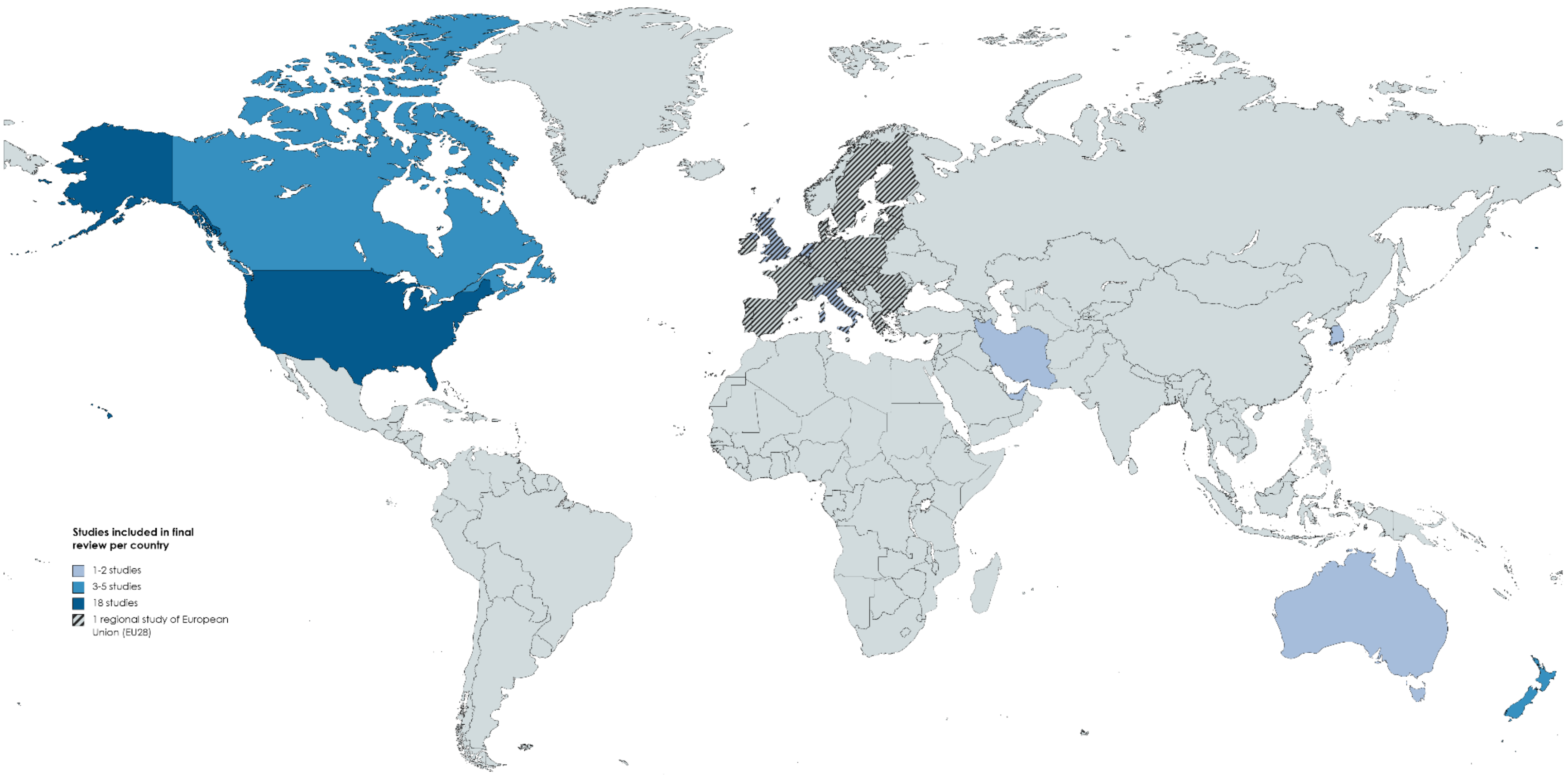


**Supplementary Figure S3**. Global distribution of studies included in final review
